# The tryptophan catabolite or kynurenine pathway in schizophrenia: meta-analysis reveals dissociations between central, serum and plasma compartments

**DOI:** 10.1101/2021.12.16.21267905

**Authors:** Abbas F. Almulla, Asara Vasupanrajit, Chavit Tunvirachaisakul, Hussein K. Al-Hakeim, Marco Solmi, Robert Verkerk, Michael Maes

## Abstract

The tryptophan catabolite (TRYCAT) pathway is implicated in the pathophysiology of schizophrenia (SCZ) since the rate-limiting enzyme indoleamine-dioxygenase (IDO) may be induced by inflammatory and oxidative stress mediators. This systematic review searched PubMed, Web of Science, and Google Scholar for papers published from inception until August 2021 and meta-analyzed the association between SCZ and TRYCATs in the central nervous system (CNS) and peripheral blood. We included 61 studies comprising 2813 patients and 2948 healthy controls. In the CNS we found a significant (p<0.001) increase in the kynurenine/tryptophan (KYN/TRP) (standardized mean difference, SMD=0.769, 95% confidence interval, CI: 0.456; 1.082) and kynurenic acid (KA)/KYN+TRP (SMD=0.697, CI:0.478-0.917) ratios, KA (SMD=0.646, CI: 0.422; 0.909) and KYN (SMD=1.238; CI: 0.590; 1.886), while the 3OH-kynurenine (3HK) + KYN-3-monooxygenase (KMO)/KYN ratio was significantly reduced (SMD=-1.089, CI: -1.682; -0.496). There were significant differences between KYN/TRP, (KYN+KA)/TRP, (3HK+KMO)/KYN, KA, and KYN levels among the CNS and peripheral blood, and among serum and plasma KYN. The only useful peripheral marker of CNS TRYCATs findings was the increased KYN/TRP ratio in serum (SMD=0.211, CI: 0.056; 0.366, p=0.007), but not in plasma. There was no significant increase in a neurotoxic composite score based on KYN, 3HK, and picolinic, xanthurenic, and quinolinic acid. SCZ is accompanied by **i**ncreased IDO activity in the CNS and serum, and reduced KMO activity and a shift towards KA production in the CNS. This CNS TRYCATs profile indicates neuroprotective, negative immunoregulatory and anti-inflammatory effects. Peripheral blood levels of TRYCATs are dissociated from CNS findings except for a modest increase in serum IDO activity.

## Introduction

Smith and Maes (1995) proposed a new neuro-inflammatory theory of schizophrenia (SCZ) that combined immune-induced neurodevelopmental abnormalities with second immune injuries, resulting in activation of immune-inflammatory and oxidative and nitrosative (O&NS) pathways, which stimulate indoleamine-dioxygenase (IDO) and the tryptophan catabolite (TRYCAT) pathway ^1^. IDO activation may result in the breakdown of TRP and an increase in the levels of different TRYCATs ^1, 2^. This early theory was updated in 2020: various SCZ phenotypes are associated not only with an activated immune-inflammatory response system (IRS), as evidenced by increased M1 macrophage activity (elevated levels of interleukin (IL)-6 and IL-1), T-helper (Th)-1, and Th-17 cells, but also with an activated compensatory immune-regulatory system (CIRS), which dampens the immune response and prevents hyperinflammation, as evidenced by increased Th-2 and T regulatory (Treg) phenotypes (e.g. increased IL-10) ^3–9^.

Increased levels of Th-1 (interferon-γ, IFN-γ) and M1 (IL-1β) cytokines, lipopolysaccharides (LPS), as well as superoxide and reactive oxygen species (ROS), may stimulate the activity of IDO, the first and rate-limiting enzyme in the TRYCAT pathway ^10–12^. Reduced TRP is a component of the innate CIRS system because reduced TRP levels protect against invading microorganisms and activation of the TRYCAT pathway plays a role in immunological tolerance and has strong antioxidant and immunosuppressive effects ^13, 14^. Furthermore, most TRYCATs (e.g. KYN, KA, XA, QA) have anti-inflammatory activities by lowering the Th-1/Treg ratio ^15^ and certain TRYCATs, especially 3-hydroxyanthranilic acid (3HA) and KA, have neuroprotective properties ^12, 16^. Furthermore, some TRYCATs are potent antioxidants, whereas some also have pro-oxidant characteristics ^11, 15^. Nonetheless, TRYCATs in the neurotoxic branch of this pathway, such as KYN, 3HK, PA, QA, and XA have behavioral and neurotoxic characteristics when overproduced ^14, 17^.

Previous research has revealed a link between TRP in the CNS and either free (i.e. unbound to albumin) or total (i.e. free and bound to albumin) plasma/serum TRP ^18, 19^. Because the competitive amino acids (CAA) leucine, isoleucine, valine, tyrosine, and phenyl alanine compete with TRP for transport through the blood brain barrier (BBB) via the large neutral amino acid transporter-1 (LAT-1), the brain levels of TRP are also governed by those CAA ^18, 19^. Furthermore, KYN and 3HK are carried to the brain at a substantial rate via LAT-1, and anthranilic acid (AA) is passively transported to the brain at a significant rate, whereas 3HA, KA, and QA have significantly lower passive diffusion rates ^20^. In chronic kidney disease or renal insufficiency, KYN and QA may cumulate in the serum and also in the cerebro-spinal fluid (CSF) as a result of increased KYN and QA transfer across the BBB and QA production in the brain ^21, 22^. Moreover, Kita et al. (2002) detected that peripheral blood KYN and QA determine in part brain KYN and QA concentrations ^23^. To further complicate matters, KYN and TRP use the same transport system to cross the BBB ^23^. It is estimated that around 60% of the KYN concentrations in the brain may be from peripheral origin ^24^.

Overall, it appears that peripheral IRS activation, particularly M1 and Th-1 activation, may result in IDO activation in the periphery and CNS, and that peripheral TRP and TRYCAT levels in part influence brain TRP, KYN, and QA levels. The several parameters that influence the availability of plasma/serum TRP/TRYCAT levels to the brain are depicted in **Figure 1**.

**Figure 1:**
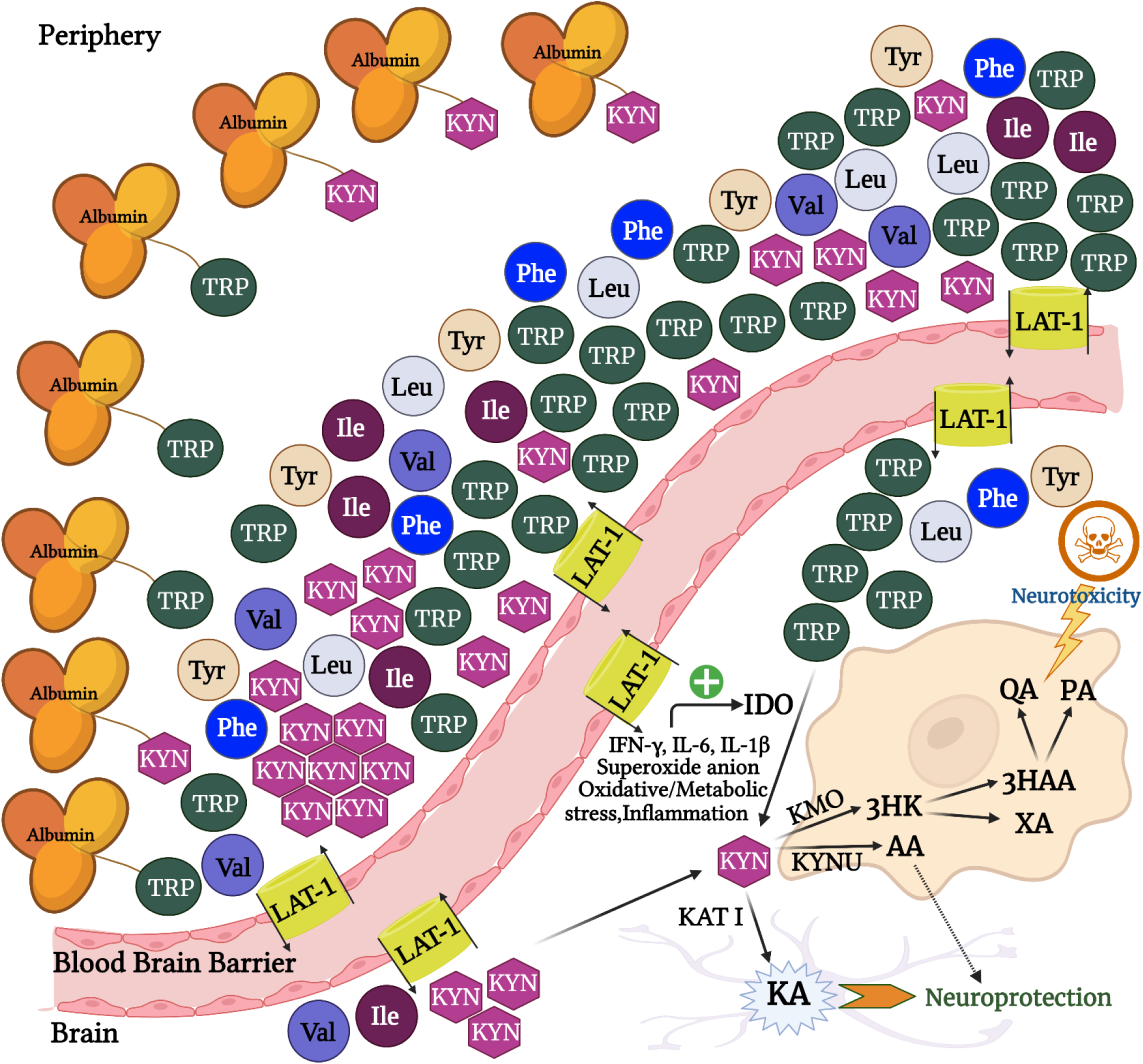
The relationship of essential amino acids and kynurenine in peripheral blood and brain tissues. This figure illustrates the transfer of tryptophan and kynurenine across the blood-brain barrier through the large neutral amino acid’s transporter-1 (LAT-1). TRP: Tryptophan, KYN: Kynurenine, Leu: Leucine, Ile: Isoleucine, Val: Valine, Phe: Phenylalanine, Tyr: Tyrosine, AA: Anthranilic acid, XA: Xanthurenic acid, 3HK: 3-Hydroxykynurenine, 3HAA: 3-Hydroxyanthranilic acid, QA: Quinolinic acid, PA: Picolinic acid, KA: Kynurenic acid, LAT-1: large neutral amino acid transporter 1, IFN-γ: Interferon-gamma, IL-6: Interleukin-6, IL-1β: Interleukin-1 beta, KMO: Kynurenine 3-monooxygenase, KATI: Kynurenine aminotransferase I, KYNU: Kynureninase, IDO: Indoleamine 2, 3-dioxygenase.

TRYCAT pathway assays in the CNS and serum/plasma of SCZ patients indicate that changes in this pathway may contribute to its pathophysiology. Firstly, elevated levels of KYN and 3HK were observed in the brain tissues and CSF of SCZ patients ^25, 26^, implying that perhaps IDO activity is increased in the brain. Another study discovered decreased levels of KMO and increased levels of KYN aminotransferase (KAT), which catalyzes the permanent conversion of KYN into KA in the brains of SCZ patients ^27^. These findings suggest a) an increased KA (neuroprotective) versus KYN (neurotoxic) ratio ^28^, and b) decreased levels of the upstream neurotoxic TRYCATs, 3HK and QA, due to decreased KMO activity ^29^. Recently, four meta-analyses examined TRP and TRYCATs levels in serum/plasma in SCZ ^30–33^. Plitman et al. investigates KA levels only and reported high KA levels in the CNS. Morrens et al. (2020) reported that some TRYCATs were downregulated in schizophrenia spectrum disorders, especially in the acute phase of the disease and older patients. Cao et al. (2021) reported that SCZ is accompanied by reduced TRP levels and a higher KYN/TRP ratio, indicating IDO activation ^32^ and significant group effects with differences in some TRYCATs between serum and plasma ^32^. Marx et al. (2021) confirmed the increased KYN/TRP ratio and reported no significant shift towards production of neurotoxic TRYCATs in SCZ ^33^.

As such, the results on the TRYCAT pathway in SCZ are contradictory and do not reveal a uniform TRYCAT pattern. Hence, the present study aimed to examine whether IDO, KMO, KAT and the neurotoxic potential of TRYCATs are elevated in the central nervous system (CNS) (brain tissues and CSF) and peripheral blood (serum and plasma) of SCZ patients versus controls. Towards this end, we conducted a systematic review and meta-analysis on the TRYCATs and TRP data. We examined the KYN/TRP and (KYN+KA)/TRP ratios (IDO proxies), (KA+KAT)/(KYN+TRP) and (KA+KAT)/KYN (KAT proxies), and (3HK+KMO)/KYN (KMO proxy) ratios or sum of all neurotoxic TRYCATs (XA, PA QA, KYN, 3HK). The specific hypotheses of the current meta-analysis are that IDO is activated, with increased levels of neurotoxic TRYCATs and lowered levels of TRP and this both in the CNS and peripheral blood.

## Materials and methods

Preferred Reporting Items for Systematic Reviews and Meta-Analyses (PRISMA) 2020 ^34^, the guidelines of the Cochrane Handbook for Systematic Reviews and Interventions ^35^, as well as the Meta-Analyses of Observational Studies in Epidemiology (MOOSE), were followed as standards guidelines to accomplished the methodology of the study. This meta-analysis investigates different TRYCATs profiles, which reflect IDO, KAT and KMO activity and the neurotoxic potential in the serum, plasma, CSF, and brain tissues, which are involved in the pathophysiology of SCZ namely prefrontal cortex^36^ and frontal cortex^26^. The KYN/TRP, (KYN+KA)/TRP, (KA+KAT)/KYN, (KA+KAT)/(KYN+TRP), and (3HK+KMO)/KYN ratios along with TRP, KYN, KA, and anthranilic acid (AA) were examined. During all stages of this study, there was no direct or indirect participation of the population or patients’ representatives.

## Search strategy

On August 11th, 2021, we began exploring the data in preparation for a systematic review, and the required data collection concluded on 15th October 2021. All related articles published within the electronic databases PubMed/MEDLINE, Google Scholar, and Web of Science were searched using precise keywords and mesh terms for the different databases to extract TRYCATs data in SCZ or schizophrenia spectrum disorder patients. All the terms and the number of identified articles is shown in the Electronic Supplementary file (ESF) Table 1. In addition, we manually checked the reference lists of the identified studies and the previous meta-analysis to ensure all published studies were included in our study.

**Table 1.** The outcomes and number of patients and controls along with the side of standardized mean difference (SMD) and the 95% confidence intervals with respect to zero SMD

| Outcome profiles | n studies | Side of 95% confidence intervals |  |  |  | Patient Cases | Control Cases | Total number of participants |
| --- | --- | --- | --- | --- | --- | --- | --- | --- |
|  |  | < 0 | Overlap 0 and SMD < 0 | Overlap 0 and SMD > 0 | > 0 |  |  |  |
| KYN/TRP | 52 | 3 | 16 | 21 | 12 | 2244 | 2483 | 4727 |
| (KYN+KA)/TRP | 61 | 6 | 20 | 21 | 14 | 2772 | 2855 | 5627 |
| (KA+KAT)/(KYN+TRP) | 62 | 8 | 18 | 22 | 14 | 2806 | 2890 | 5696 |
| KA | 34 | 8 | 7 | 9 | 10 | 1615 | 1659 | 3274 |
| TRP | 44 | 14 | 19 | 7 | 4 | 1921 | 2181 | 4102 |
| KYN | 37 | 6 | 14 | 9 | 8 | 1670 | 1865 | 3535 |
| (KA+KAT)/KYN | 49 | 7 | 18 | 15 | 9 | 2367 | 2386 | 4753 |
| (3HK+KMO)/KYN | 41 | 11 | 12 | 13 | 5 | 1860 | 2036 | 3896 |
| (KYN+3HK+PA+QA+XA) | 44 | 10 | 16 | 12 | 6 | 2042 | 2151 | 4193 |

KYN: Kynurenine, TRP: Tryptophan, KA: Kynurenic acid, 3HK: 3-Hydroxykynurenine, PA: Picolinic acid, QA: Quinolinic acid, XA: Xanthurenic acid, KMO: Kynurenine 3-monooxygenase, KAT: Kynurenine aminotransferase.

## Eligibility criteria

The English language and publication in peer-reviewed journals were the primary criteria for including manuscripts, but grey literature, papers in other languages (Thai, French, Spanish, German, Italian, Arabic) and the reference lists of the extracted papers were also examined. Inclusion criteria comprise a) observational case-control and cohort studies which investigate the concentrations of TRYCATs (and TRP in the same studies) in serum, plasma, CSF, and brain tissues of the patients with SCZ or schizophrenia spectrum disorders; b) the patients should be diagnosed according to any version of the Diagnostic and Statistical Manual of Mental Disorders (DSM) or the International Classification of Diseases (ICD) criteria as SCZ or psychotic spectrum disorders; and c) studies that include a normal control group. Exclusion criteria were: a) studies conducted on animal samples and genetic and translational studies; b) lack of a normal control group; c) systematic reviews or meta-analyses; d) articles with duplicate data, and e) studies not reporting the means and standard deviation (SD)/standard error (SEM) values, and f) studies that utilized saliva or whole blood to assess TRYCATs. Nevertheless, we contacted the authors of studies that did not show mean with SD/SEM values but showed geometric means, median (interquartile range) or graph format data. When the author did not respond, and the median with either interquartile range (IQR) or minimum/maximum values was presented, we estimated the mean and SD values according to a method suggested by ^37^ , or we used the Web Plot Digitizer to estimate mean and SD/SEM from graphs (https://automeris.io/WebPlotDigitizer/).

### Primary and secondary outcomes

The primary outcome measures of the present study are the KYN/TRP and (KYN+KA)/TRP ratios, reflecting IDO activity ^28^, in patients with SCZ versus controls (see **Table 1**). Secondary outcome variables were (KA+KAT)/KYN and (KA+KAT)/(KYN+TRP) (KAT activity proxies), and (3HK+KMO)/KYN ratio (KMO activity proxy). In addition, if there were any differences in these ratios, we also conduct meta-analyses on the solitary TRYCATs (KYN, 3HK, KA, AA) and TRP. The meta-analysis estimated the neurotoxic potential of the produced TRYCATs by entering a composite score based on KYN, 3HK, XA, QA, and PA, and examining the (KA+KAT)/KYN ratio.

### Screening and data extraction

The first author (AA) conducted a preliminary review of the studies to determine whether each study could be included in the current meta-analysis based on the inclusion criteria by inspecting the titles and abstracts of the papers. After eliminating studies based on the pre-defined exclusion criteria, the entire text of potentially eligible publications was downloaded. The same author also extracted the mean and SD, and other required data from the selected articles. They were first organized on a pre-defined excel spreadsheet designed specifically for this purpose. Once the first author (AA) had completed all information, another author (AV) double-checked the retrieved data. In the case of a conflict, the last author (MM) was consulted.

The key data in the pre-defined excel file comprised: author’s name, year of publication, names of measured TRYCATs, mean and SD of the biomarkers, as well as the sample size for each group (patients and healthy controls, type of the study, demographic characteristics of the study population involved (mean±SD of age and sex distribution), rating scales for determining the severity of the disease, the medium, which was examined to measure the TRYCATs (brain tissue, CSF, serum, and plasma) as well the study’s latitude and scores of methodological quality. The immune cofounder scale (ICS) ^38^ was utilized as a methodological quality score checklist, which the last author somewhat adjusted to be useful for TRYCATs data. Both scoring scale (quality and redpoint) checklists used in the present meta-analysis are shown in ESF Table 2. The major purpose of using such score scales is to evaluate the methodological quality of studies conducted to measure the TRYCATs concentrations. The first scale focuses on key quality domains of the articles consisting of sample size, confounder control, the exact time for collecting the samples, etc. The total scores range from 0 to 10, with values to ten scores denoting higher methodological quality. The second scale is the redpoints score scale which assesses the lack of control or adjustment for key confounders and thus increased analytical/biological bias in the TRYCATs assays and study designs. Total scores can vary from 0 to 26, with 0 indicating that all confounder variables were considered and 26 indicating that there was no control at all.

**Table 2.** Results of meta-analysis performed on several outcome (TRYCATs) variables and different media, central nervous system (CNS), serum, and plasma, alone and together.

| Outcome feature sets | n | Groups | SMD | 95% CI | z | p | Q | df | p | I<sup>2</sup> (%) | $\tau^2$ | T |
| --- | --- | --- | --- | --- | --- | --- | --- | --- | --- | --- | --- | --- |
| KYN/TRP | 52 | Overall | 0.213 | 0.105; 0.321 | 3.86 | < 0.0001 | 170.90 | 51 | < 0.0001 | 70.15 | 0.117 | 0.343 |
|  | 5 | CNS | 0.769 | 0.456; 1.082 | 4.82 | < 0.0001 | 5.16 | 4 | 0.272 | 22.44 | 0.029 | 0.169 |
|  | 21 | Serum | 0.211 | 0.056; 0.366 | 2.68 | 0.007 | 42.16 | 20 | 0.003 | 52.57 | 0.058 | 0.240 |
|  | 26 | Plasma | 0.046 | -0.126; 0.218 | 0.52 | 0.600 | 97.16 | 25 | < 0.0001 | 74.27 | 0.133 | 0.365 |
| (KYN+KA)/TRP | 61 | Overall | 0.177 | 0.069; 0.285 | 3.20 | 0.001 | 243.94 | 60 | < 0.0001 | 75.40 | 0.153 | 0.391 |
|  | 10 | CNS | 0.714 | 0.465; 0.963 | 5.63 | <0.0001 | 15.33 | 9 | 0.082 | 41.30 | 0.063 | 0.252 |
|  | 22 | Serum | 0.167 | -0.034; 0.367 | 1.63 | 0.103 | 84.80 | 21 | < 0.0001 | 75.24 | 0.148 | 0.385 |
|  | 29 | Plasma | -0.013 | -0.163; 0.137 | -0.17 | 0.866 | 94.37 | 28 | < 0.0001 | 70.33 | 0.108 | 0.328 |
| (KA+KAT)/(KYN+TRP) | 62 | Overall | 0.150 | 0.026; 0.274 | 2.37 | 0.018 | 273.23 | 61 | < 0.0001 | 77.67 | 0.174 | 0.417 |
| (KA+KAT)/KYN | 49 | Overall | 0.072 | -0.069; 0.212 | 0.99 | 0.320 | 236.21 | 48 | < 0.0001 | 79.68 | 0.184 | 0.429 |
| (3HK+KMO)/KYN | 41 | Overall | 0.023 | -0.109; 0.155 | 0.33 | 0.735 | 231.93 | 40 | < 0.0001 | 82.75 | 0.225 | 0.474 |
|  | 6 | CNS | -1.089 | -1.682; -0.496 | -3.59 | <0.0001 | 28.40 | 5 | < 0.0001 | 82.40 | 0.444 | 0.666 |
|  | 16 | Serum | -0.182 | -0.441; 0.077 | -1.37 | 0.169 | 67.11 | 15 | < 0.0001 | 77.65 | 0.187 | 0.433 |
|  | 19 | Plasma | 0.179 | 0.021; 0.338 | 2.21 | 0.027 | 52.87 | 18 | < 0.0001 | 65.95 | 0.075 | 0.274 |
| KA | 34 | Overall | 0.288 | 0.119; 0.456 | 3.34 | 0.001 | 332.43 | 33 | < 0.0001 | 90.07 | 0.436 | 0.661 |
|  | 10 | CNS | 0.676 | 0.442; 0.909 | 5.67 | <0.0001 | 13.78 | 9 | 0.130 | 34.68 | 0.047 | 0.217 |
|  | 14 | Serum | 0.042 | -0.380; 0.464 | 0.19 | 0.846 | 192.61 | 13 | < 0.0001 | 93.25 | 0.562 | 0.750 |
|  | 10 | Plasma | -0.225 | -0.524; 0.074 | -1.47 | 0.141 | 46.96 | 9 | < 0.0001 | 80.83 | 0.180 | 0.425 |
| TRP | 44 | Overall | -0.213 | -0.364; -0.088 | -3.21 | 0.001 | 194.32 | 43 | < 0.0001 | 77.87 | 0.173 | 0.416 |
| KYN | 37 | Overall | -0.107 | -0.235; 0.021 | -1.64 | 0.101 | 216.13 | 36 | < 0.0001 | 83.34 | 0.236 | 0.486 |
|  | 5 | CNS | 1.238 | 0.590; 1.886 | 3.74 | <0.0001 | 20.29 | 4 | < 0.0001 | 80.29 | 0.432 | 0.657 |
|  | 16 | Serum | 0.152 | -0.111; 0.416 | 1.13 | 0.257 | 69.85 | 15 | < 0.0001 | 78.53 | 0.197 | 0.443 |
|  | 16 | Plasma | -0.263 | -0.414; -0.113 | -3.44 | 0.001 | 32.55 | 15 | 0.005 | 53.91 | 0.045 | 0.212 |
| (KYN+3HK+PA QA+XA) | 44 | Overall | -0.052 | -0.198; 0.093 | -0.70 | 0.481 | 255.75 | 43 | < 0.0001 | 83.19 | 0.237 | 0.487 |

KYN: Kynurenine, TRP: Tryptophan, KA: Kynurenic acid, 3HK: 3-Hydroxykynurenine, PA: Picolinic acid, QA: Quinolinic acid, XA: Xanthurenic acid, KAT: Kynurenine aminotransferase, KMO: Kynurenine 3-monooxygenase, CNS: Brain and CSF, SMD: standardized mean difference, 95% CI: 95% confidence intervals

### Data analysis

The CMA V3 program was used to conduct a PRISMA-based meta-analysis. **Table 1** shows the outcome biomarker profiles examined in our systematic review and meta-analysis. Meta-analyses were conducted for variables that were presented in at least three studies. The synthetic scores reflecting IDO, KMO and KAT activity and neurotoxic potential were compared among SCZ patients and healthy controls by computing the mean values of the markers of their respective profiles while assuming dependence. To estimate IDO activity, KYN and TRP were entered in the analysis with the direction of increasing KYN and decreasing TRP favoring SCZ. The (KA+KAT)/KYN or (KA+KAT)/(KYN+TRP) ratios (estimating KAT) were estimated by entering KA and KAT (direction set at positive, favoring SCZ), and KYN and TRP (direction: negative, favoring SCZ) were entered in the meta-analysis. The (3HK+KMO)/KYN ratio was estimated by entering 3HK and KMO, direction positive, favoring SCZ, and KYN, direction set at negative, favoring SCZ, in the analysis. The total neurotoxic potential of the TRYCAT pathway was estimated by entering all neurotoxic TRYCATs (XA, PA, KYN, 3HK and QA) in the meta-analysis.

We conducted the meta-analysis employing a restricted maximum-likelihood random-effects model under the assumption that the features of the included studies varied. The effect size was estimated by computing the standardized mean difference (SMD) with 95% confidence intervals (95% CI). The results were statistically significant at p <0.05 (two-tailed tests). SMD values of 0.2, 0.5 and 0.8 indicate small, moderate, and large effect sizes, respectively ^39^. We used tau-squared values to denote heterogeneity, as previously described, and we also calculated Q and I^2^ metrics ^40, 41^. This meta-analysis examined subgroups (brain tissue, CSF, serum, plasma) and used those subgroups within the study as unit of analysis. Brain tissue and CFS were combined into a “central nervous system (CNS)” subgroup if there were no significant differences in the outcome variables between these groups. The meta-analysis was also performed across the subgroup levels and we compared the effects at the different levels within the study.

## Results

### Search results

We examined 2025 research papers according to the specific keywords and search sentences listed in ESF, table 1 during the search process. **Figure 2** shows the Prisma flow chart and displays the search method’s outcomes and details for inclusion-exclusion articles. After removing 1957 records from the initial search outcome, 68 full-text research papers were eligible for the systematic review. Seven of the 68 articles were eliminated for the reasons listed in ESF, table 4 and therefore the meta-analysis consists of 61 records ^25–27, 36, 42–98^.

**Figure 2:**
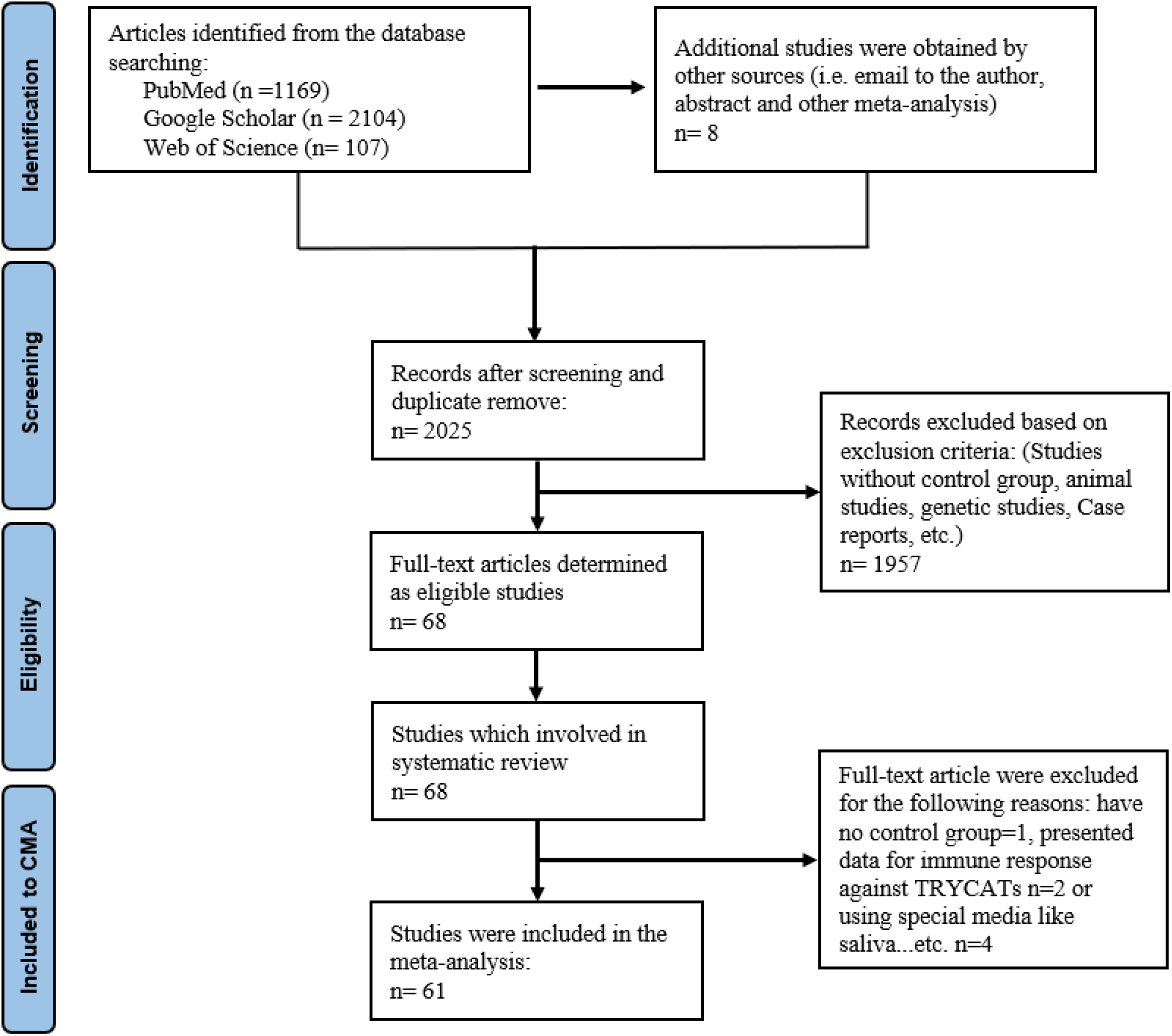
Prisma flow-chart of the meta-analysis study of tryptophan catabolites in schizophrenia

The total number of participants included in the current meta-analysis is 5761, namely 2813 SCZ patients and 2948 healthy controls. This systematic review and meta-analysis only included case-control studies. Seven studies were excluded from the meta-analysis and ESF, table 4 shows the reasons. Twelve studies assayed the TRYCAT pathway in the CNS (8 CSF and 4 brain tissue studies), 27 studies in the plasma, and 22 in the serum. The included papers investigated the TRYCAT pathway in the CNS of 677 individuals (318 patients versus 359 healthy controls), serum of 2186 participants (1054 patients versus 1132 healthy controls), and plasma of 2899 subjects (1441 patients and 1458 healthy subjects). The most common method for assaying TRYCATs was high-performance liquid chromatography (HPLC) used in 31 studies. In 19 research papers, liquid chromatography-mass spectrometry (LCMS) was utilized, while other studies used alternative test procedures indicated in ESF, table 5.

Overall, there were 12 studies conducted on SCZ spectrum disorder patients and 49 studies on pure SCZ patients. We included 2065 SCZ patients versus 2005 healthy controls and 747 patients with SCZ spectrum disorders versus 943 healthy controls. The participants’ ages ranged from 20 to 60 years old. Most of the participants in the current meta-analysis are from the United States (19 studies), Sweden (8 studies), China (8 studies), Japan (4 studies), Italy (3 studies), Netherlands (3 studies), Brazil (2 studies), South Korea (2 studies), Germany (2 studies), Australia, Austria, Belgium, Estonia, India, Ireland, Norway, Poland, Switzerland, UK (each one). The quality scores along with redpoint scores, are shown in ESF, table 5 as median (min-mix), namely 4 (min= 1, max=8) and 12 (min=7.0, max=25.0), respectively.

### The primary outcome variables

#### KYN/TRP

**Table 1** shows that in 12 studies (4 CNS, 3 plasma, 5 serum studies), the 95% CI intervals of KYN/TRP were totally on the positive side of zero while 3 studies (2 plasma, 1 serum) reported 95% CI which were entirely on the negative side of zero. Additionally, 36 intervals crossed the zero distributed as 21 studies with SMD values greater than zero and 16 studies with SMD smaller than zero. The forest plot of KYN/TRP in patients versus controls is shown in **Figure 3**. There were no significant differences (p=0.407) in the ratio between brain tissue (SMD=0.606; 95% CI: 0.175, 1.038, p=0.006) and CSF (SMD=0.877; 95%CI: 0.406, 1.348, p=0.001) and, therefore, we examined the CNS subgroup combining CSF and brain tissues. The meta-analysis performed on 5 CNS, 26 plasma, and 21 serum studies showed a statistically significant SMD with a small effect size. However, high heterogeneity was observed when considering all media together, and, therefore, subgroup analysis was carried out. There were significant differences between the three media (p<0.0001) with a high effect size in the CNS and a small but significant effect size in serum, whereas plasma yielded non-significant results (see **Table 2**). There were significant differences among CNS and serum (p=0.002) and plasma (p<0.0001) and a trend toward a non-significant difference between plasma and serum (p=0.162). The heterogeneity in CNS and serum was lower than in the total sample and plasma. ESF, table 6 displays that no bias was present when all data were combined and the CNS, serum and plasma did not show any signs of bias.

**Figure 3:**
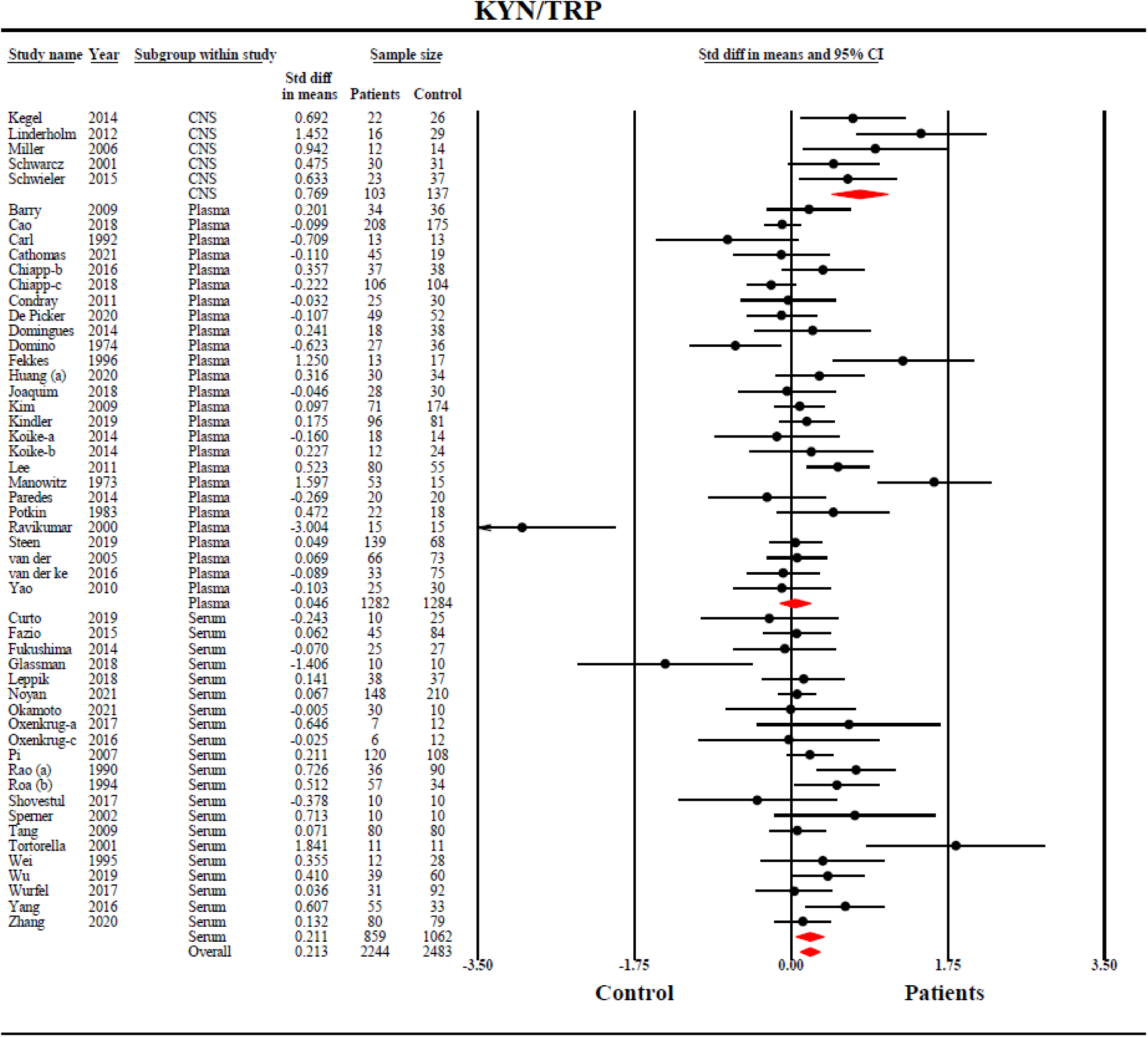
Forest plot of the kynurenine/tryptophan (KYN/TRP) ratio reflecting indoleamine-2,3-dioxygenase (IDO) activity with results of a meta-analysis conducted upon 52 studies.

#### The (KYN+KA)/TRP ratio

There were no significant differences (p=0.562) in the (KYN+KA)/TRP ratio between brain tissue (SMD=0.890; 95% CI: 0.177, 1.603, p=0.014) and CSF (SMD=0.665; 95% CI: 0.444, 0.940, p<0.001). **Table 1** shows that fourteen studies (6 CNS, 3 plasma, 5 serum) showed 95% CI that were totally on the positive side of zero while 6 studies (4 plasma, 2 serum) reported CI values that were entirely on the negative side of zero. Additionally, 41 intervals crossed the zero, distributed as 21 studies with SMD values greater than zero and 20 studies with SMD smaller than zero. ESF, Figure 1 shows the forest plot of the (KYN+KA)/TRP ratio. The results indicate an overall significant SMD of 0.177, although heterogeneity was considerable and again partly explained by highly significant intergroup differences (p<0.0001) with a high effect size in the CNS and no significant effects sizes in serum and plasma (see **Table 2**). There were highly significant differences between CNS and serum (p=0.001) and plasma (p<0.0001). As shown in ESF, table 6, publication bias occurs when all data are combined, resulting in a decrease in the adjusted point estimate (0.132; 95% CI: 0.008; 0.255). Significant bias was detected in plasma as displayed in ESF, table 6, and the adjusted point estimate was 0.05 with 95% CI (-0.104; 0.205). In contrast, there was no bias when considering CNS and serum.

### Secondary outcome variables

#### (KA+KAT)/(KYN+TRP) ratio

There were no significant differences (p=0.530) in the (KA+KAT)/(KYN+TRP) ratio between brain tissue (SMD=0.503; 95% CI: -0.217, 1.222, p=0.171) and CSF (SMD=0.246; 95% CI: -0.106, 0.897, p=0.171). **Table 2** shows that the (KA+KAT)/(KYN+TRP) ratio was significantly higher in SCZ than in controls with a very modest effect size. There were no significant differences between CNS, serum, and plasma (p=0.463). Nevertheless, the ratio in CNS was significantly higher in SCZ as compared with controls, with a moderate effect size (SMD=0.325, 95% CI: 0.010; 0.639, tau^2^=0.184), whereas the effects sizes were not significant in serum (SMD=0.082, 95% CI: -0.153; 0.317, tau^2^=0.229) and plasma (SMD=0.136, 95% CI: -0.030; 0.301, tau^2^=0.142). ESF, Figure 2 shows the forest plot of the (KA+KAT)/(KYN+TRP) ratio and ESF, table 6 shows that probably no bias was present.

#### (KA+KAT)/KYN

There were no significant differences (p=0.642) in the (KA+KAT)/KYN ratio between brain tissue (SMD=0.446; 95% CI: -0.630, 1.521, p=0.417) and CSF (SMD=0.172; 95% CI: - 0.245, 0.589, p=0.419). The results displayed that overall, there was no significant difference in the (KA+KAT)/KYN ratio between SCZ patients and controls (SMD=0.072, 95% CI: -0.069; 0.212, tau^2^= 0.184) and that no significant differences were detected between CNS, serum and plasma (p=0.217). After adjusting for 2 missing studies the overall SMD was 0.091 (95% CI: - 0.050; 0.232).

#### (3HK+KMO)/KYN

**Table 2** shows that the (3HK+KMO)/KYN ratio was not significantly different between SCZ patients and controls. Since there were highly significant differences between CNS, serum, and plasma (p<0.0001) we performed subgroup analysis and found that the CNS (3HK+KMO)/KYN ratio was significantly lower in SCZ than in controls (SMD=-1.089, 95%CI: -1.682; -0.496, tau^2^= 0.444), whereas plasma showed an increased ratio in SCZ (SMD= 0.179, 95%CI: 0.021; 0.338, tau2= 0.075). There were highly significant differences between CNS and plasma (p<0.0001) and CNS and serum (p=0.006). In plasma, imputation of 6 studies changed the SMD to 0.323 (95% CI: 0.167; 0.479).

#### TRP, KYN, KA and AA

**Table 2** and ESF, Figure 3 show that TRP was significantly lower in SCZ patients than in controls with a very modest effect size. There were no significant differences in the TRP effect sizes among serum, plasma, and CSF. Kendall’s tau and Egger’s regression did not suggest that bias could be present. Nevertheless, after imputing three missing studies on the right site, the adjusted estimated point estimate (-0.081) was no longer significant (95% CI: -0.241; 0.078).

**Table 2** shows that in all studies combined there was no significant change in KYN in SCZ versus controls. Nevertheless, group analysis showed highly significant differences between CNS, serum and plasma (p<0.0001) with significant differences between CNS and either serum (p=0.002) and plasma (p<0.0001) and between serum and plasma (p=0.007). While in the CNS (high effect size) and serum (non-significant) there was a positive association with SCZ, in plasma a highly significant inverse correlation was established (see **Table 2**). **Figure 4** shows the forest plot of the KYN data in SCZ. Kendall’s tau and Egger’s regression did not suggest that bias was present in CNS, serum, or plasma.

**Figure 4.**
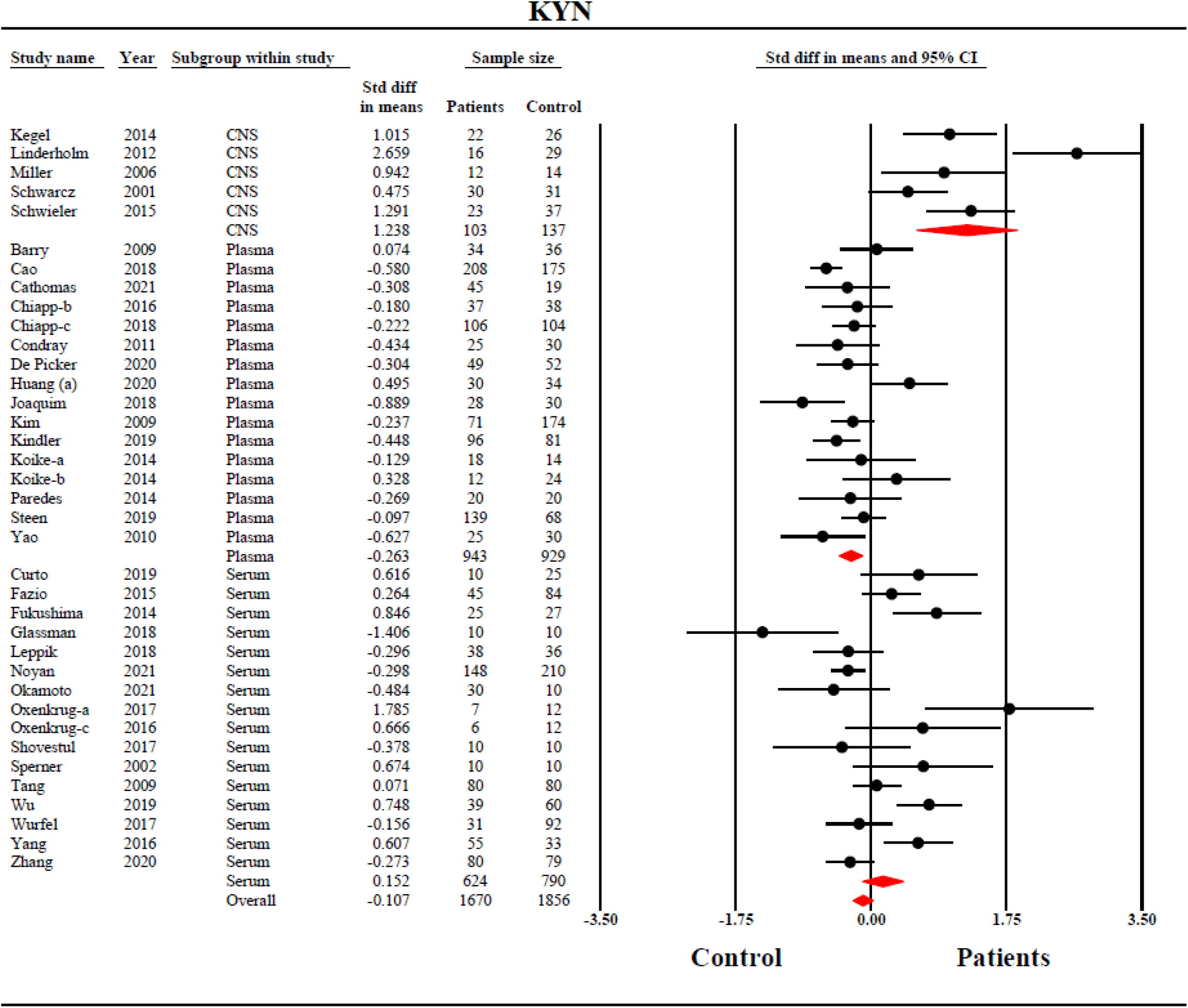
Forest plot with results of the meta-analysis performed on 37 studies reporting on kynurenine (KYN) in schizophrenia.

**Table 2** and ESF, Figure 4 show that KA was significantly higher in all SCZ patients combined than in controls with a very modest effect size. Group analysis showed highly significant differences between CNS, serum, and plasma (p<0.0001) with significant differences between CNS and either plasma (p<0.0001) and serum (p=0.01). ESF, table 6 shows the results on publication bias in the KA data which show a bias in the plasma but not serum levels. Anthranilic acid (AA) data were obtained in three serum studies and show significantly increased AA levels in SCZ as compared with controls with a medium effect size (SMD= 0.590, 95% CI: 0.045; 1.136, p=0.034).

#### (KYN+3HK+PA+QA+XA) composite

The composite score of KYN,3HK, PA, QA and XA was not significant different between SCZ patients and controls. In addition, group analysis showed no significant difference (p=0.344) among CNS, serum, and plasma. Publication bias was detected with 4 missing studies and the adjusted SMD was 0.134 (95% CI: -0.048; 0.318).

## Meta-regression analyses

Meta-regression (ESF, table 7) reveals that after adjusting for the differences between CNS, serum and plasma, there were significant effects of latitude (p=0.032) on the KYN/TRP ratio and KYN. Age explained part of heterogeneity in the KYN/TRP ratio and KYN. Female gender affects the results of (KYN+KA)/TRP ratio, KA and KYN. The total number of participants affected heterogeneity in KYN.

## Discussion

### The KYN/TRP ratio in SCZ

The current study’s first major finding is that the KYN/TRP ratio was significantly increased in SCZ patients when compared to controls, implying that IDO may be activated in SCZ patients. Nonetheless, subgroup analysis across the CNS, serum, and plasma reveals a large effect size (0.769) in the CNS and a small effect size (0.211) in the serum, whereas the effect size in the plasma is non-significant. As such, serum measurements reflect changes in the CNS in part, whereas plasma measurements are completely unrelated to CNS observations. A such, the current study discovered a more widespread IDO activation in the CNS and serum of SCZ patients. It appears that the plasma KYN/TRP ratio cannot be used as a proxy for IDO activity in the brain or peripheral blood. Moreover, computing the (KYN+KA)/TRP ratio showed significant differences in CNS but not in serum, suggesting that it is more adequate to interpret the serum KYN/TRP ratio when evaluating peripheral blood IDO activity. The findings are in agreement with the theories (see Introduction) that activated M1, Th-1, and O&NS pathways in SCZ stimulate IDO, resulting in TRYCAT pathway activation ^99–102^.

Secondary analyses to determine whether KYN or TRP or both are altered in SCZ revealed that KYN levels were significantly increased in SCZ patients compared to controls, and that there were highly significant differences between CNS and serum or plasma and between plasma and serum. Thus, KYN levels were significantly increased in the CNS of SCZ patients and slightly increased in the serum, whereas the plasma assay revealed a significant decrease in KYN. As a result, the findings in plasma are diametrically opposed to those in the CNS. Morrens et al. ^31^, on the other hand, did not account for plasma-serum differences, and thus their KYN and KYN/TRP results are uninterpretable. Two other meta-analyses also detected increased serum KYN but decreased plasma KYN levels in SCZ ^32 33^. Nonetheless, the primary distinction between our study and those two meta-analyses is that we calculated the mean of all available data on KYN and TRP in all studies as well as a composite score for the KYN/TRP ratio ^40, 103^. On the other hand, Cao et al. and Marx et al. used only a few KYN/TRP ratios when the original articles computed this ratio. Consequently, the current meta-analysis findings far surpass those of the earlier studies.

Group analysis performed on the TRP levels, on the other hand, did not show significant differences between CNS, plasma, and serum. Although the overall meta-analysis showed lower TRP levels in SCZ than in controls, these differences were no longer significant after imputing missing values. Three previous meta-analyses ^31–33^ reported an overall significant reduction in TRP level in SCZ.

Because KYN can cross the BBB during systemic inflammation, blood derived KYN is the primary source of brain KYN levels (see Introduction and **Figure 1**). Additionally, the brain concentrations of other TRYCATs, such as KA, 3HK, and QA, are determined in part by peripheral TRYCATs levels ^23, 104^. As such, alterations in peripheral blood TRYCATs induced by peripheral inflammation contribute to a variety of immune, redox, and behavioral effects in the brain (see Introduction).

### KAT activity in SCZ

The second major finding of this study is that the (KA+KAT)/(KYN+TRP) ratio and KA levels, but not the (KA+KAT)/KYN ratio, are significantly increased in the CNS of SCZ patients, whereas serum and plasma showed no significant effects or even an inverse association. These findings extend those of Plitman et al. (2017) and Cao et al. ^30, 32^, who discovered increased brain KA levels in SCZ but no significant changes in peripheral KA. Marx et al. ^33^ reported that SCZ patients had stable KA levels and a decreased KA/KYN ratio. Morrens et al. ^31^ found no statistically significant differences in peripheral blood KA levels between SCZ and controls, but as previously stated, these results are not interpretable. As discussed above, our findings regarding ratios, including the (KA+KAT)/KYN ratio, are much more appropriate and should be considered when interpreting those ratios.

Our findings suggest that increased KA formation from its precursors KYN and TRP may occur in the CNS, whereas no such effects were observed in the peripheral blood. Such disparities between the brain and peripheral blood have also been observed during aging, which is associated with increased KAT activity in the brain but no changes in the periphery ^105^. KA is unlikely to cross the BBB ^20^, and the majority of brain KA is derived from KYN ^104^. Thus, blood borne KYN, which is transported from the peripheral blood to the brain, may stimulate KAT activity in the CNS (see Introduction). This explains that blood-derived levels of KYN are correlated with CNS KA concentrations ^25, 26^. Astrocytes are another critical determinant of KA level in the CNS because they lack KMO (KYN hydroxylase) and produce KA and KYN in response to IRS-induced IDO activation ^106^. For instance, elevated KA levels may be detected following IL-6 administration to cultured astrocytic cells ^44^.

### KMO in SCZ

The third major finding of this meta-analysis is that the (3HK+KMO)/(KYN+TRP) ratio (a proxy for KMO activity) was significantly decreased in the CNS, whereas no significant changes were observed in plasma or serum. Not only increased IDO activity, but also decreased KMO activity may account for some of the increases in KYN and KA levels in the CNS of SCZ patients ^107, 108^. The onset of inflammation and infection are accompanied by depletion of intracellular riboflavin resulting in lowered KMO activity ^109^, while reduced KMO activity induces elevated levels of AA concentrations and an increase in the utilization of KYN, resulting in increased KA production ^109, 110^. Our meta-analysis revealed that AA levels were significantly elevated in SCZ, even though only three studies assessed AA levels. Notably, AA serves as a protective agent because this TRYCAT may inhibit the formation of QA and PA from 3HA ^109, 111^. Moreover, 3HA has many potent anti-inflammatory and negative immune-regulatory effects ^109^.

### Is there increased neurotoxicity in SCZ?

The fourth major finding of our study is that SCZ was not accompanied by a significant increase in the composite KYN + 3HK + XA + PA + QA score, a neurotoxicity index. Reduced KMO activity and increased AA concentrations may have contributed to the absence of significant increases in neurotoxic TRYCATs such as PA and QA despite increased IDO activity. Additionally, as mentioned previously, the (KA+KAT)/KYN ratio, a measure of neuroprotection/neurotoxicity, was significantly increased in SCZ, implying a net neuroprotective effect. KA appears to have more antioxidant than pro-oxidant properties, exhibiting significant ROS scavenging activity against hydroxyl, peroxinitrite, and superoxide, as well as inhibiting lipid peroxidation and subsequent aldehyde formation and protein oxidation ^105^. In addition, KA is a CIRS component that has anti-inflammatory properties and aids in the resolution of inflammation ^112^. KA also exerts neuroprotective effects ^12, 113^ by inhibiting extra synaptic NMDA-receptors and the 7 nicotinic acetylcholine receptor (7nAChr) thereby preventing the release of glutamate and thus further stimulation of postsynaptic NMDA-receptors ^114^. The preceding demonstrates that KA has a multitude of protective properties and may act as a buffer against the neurotoxic effects of QA and KYN, while the latter has also some anti-inflammatory and antioxidant properties which contribute to neuroprotection and CIRS activity ^102, 105^. These protective effects should be added to the pathway’s intrinsic functions comprising CIRS, anti-inflammatory, and antioxidant activities (see Introduction).

At first glance, the findings of this meta-analysis contradict prior reviews’ claims that SCZ is associated with greater TRYCATs neurotoxicity ^12^. However, new research using a more sensitive TRYCATs assay which measures serum IgA levels directed to TRYCATs shows that only first episode schizophrenia (FES)/multiple episode schizophrenia (MES) with worsening and consequent deficit SCZ are characterized by increased TRYCAT-associated neurotoxicity, as evidenced by the measurement of IgA levels to XA, PA, and 3HK ^115^. This IgA response evaluates changes in TRYCATs levels as identified by the immune system. According to these recent findings, elevated neurotoxic TRYCATs may play a role in the neuroimmune toxicity linked to the worsening of FES/MES and SCZ deficiency ^115^. As a result, one of the major flaws in TRYCATs studies in SCZ is that they failed to account for the worsening of MES and FES, as well as the resulting deficit SCZ ^115^.

### Sources of heterogeneity in TRYCATs studies

Apart from the major differences in TRYCATs levels between CNS, plasma and serum, subgroup analyses showed that latitude, age, gender, and the number of participants in the studies may contribute to the heterogeneity. Moreover, significantly increased KA, KYN, and KYN/TRP and (KYN+KA)/TRP ratios in patients who are treated with antipsychotics although these results may be confounded by to differences among serum, plasma and CNS. The effects of antipsychotics in modulating TRYCATs is still ambiguous.

Our findings show that the plasma assay of TRYCATs and plasma KYN may be prone to increased sources of pre-analytical or analytical error. First, the dilution effects of anticoagulants may impact lower concentrations of analytes ^116^. Furthermore, thermally decomposition following EDTA may increase α-amino products ^117^ and carbonyl-containing compounds such as EDTA may induce degradation of KYN in biological samples ^118^.

There are some studies suggesting that - to achieve the most precise TRP assay - it is preferable to collect serum instead of plasma to avoid pre-analytical and analytical errors emerging from using plasma anti-coagulant tubes based on EDTA, heparin, and citrate ^119, 120^. These materials may interfere with the metabolites thereby producing contamination especially in HPLC and spectrophotometric methods. Furthermore, the probability of oxidative degradation of TRP is high, and carbonyl-containing compounds including EDTA may contribute to TRP degradation ^118^. Moreover, at room temperature (> 4 °C), serum TRP and the CAA may increase in plasma (personal observations), which may be explained by increased protease activity which release amino acids. Moreover, also heparin tubes may not be appropriate to assay TRP as heparin may interact with binding of albumin to TRP ^121, 122^. Another source of variation in plasma/serum TRP levels is associated with blood platelets, which have high concentrations of TRP and 5-HT and may release TRP when activated. Patients with SCZ show altered platelet reactivity due to O&NS ^123^, which may impact 5-HT levels in platelets and consequently impact peripheral TRP concentrations ^124^.

Apart from the effects of the above sources of (pre-)analytical variability, the TRYCAT pathway products are also prone to considerable biological variability. First, the ratio of free or total plasma/serum TRP / the sum of the 5 CAA is probably the best index predicting brain TRP concentrations ^14^. Second, TRP is bound (80%) to albumin ^19, 125^ and, thus, any changes in albumin (a negative acute phase protein which is decreased in inflammatory conditions) are accompanied by changes in TRP levels. Moreover, TRP is stripped off from albumin in the microcirculation when transported through the BBB ^126^. Third, insulin levels indirectly effect this binding by decreasing plasma levels of non-esterified fatty acids which compete with TRP for albumin binding ^19, 127^.

## Limitations

The current systemic review and meta-analysis’ findings should be addressed in terms of their limitations. Firstly, to delineate whether TRYCAT-associated neurotoxicity (especially in PA, XA, and QA) plays a role in SCZ, we would need more brain tissue, CSF and serum levels of the neurotoxic TRYCATs including in the FES and MES worsening phenotypes and deficit SCZ. Some TRYCATs were largely missing in the studies included here, e.g. 3HA, and therefore we were unable to compute the 3HA/AA ratio which is important to understand the protective properties of the TRYCAT pathway. Third, in the present study we combined TRYCAT measurements in CFS and relevant brain tissues into a new subgroup, namely CNS, because we could not detect any differences between both compartments and to increase the number of studies to be included in the meta-analysis. Nevertheless, more studies on both compartments are needed to further examine possible differences and effects of for example the organic anion transporter which is involved in the transport of KA into CSF ^128^. Fourth, preanalytical studies should scrutinize why there are significant differences in some TRYCATs between plasma and serum. Meanwhile we recommend measuring peripheral levels of TRYCATs in serum and not plasma.

## Conclusions

**Figure 5** summarizes the findings of the current meta-analysis. Increased IDO and decreased KMO activity accompany SCZ, showing that the TRYCAT pathway’s first part is activated. This leads to increased KA production in the CNS, which improves neuroprotection, antioxidant protection, and CIRS activities, as well as enhanced AA production, which may reduce neurotoxic TRYCATs like QA and PA. SCZ does not show a substantial rise in the composite score, which includes all neurotoxic TRYCATs. Only the serum KYN/TRP ratio appears to be linked to increased KYN levels and IDO activity in the CNS. The other TRYCAT levels in the peripheral blood have no diagnostic significance. Even worse, when compared to the CNS, plasma TRYCAT levels demonstrate the opposite results. The TRYCATs profile established in SCZ patients is predominantly neuroprotective (increased KA and AA) and should be considered in addition to the pathway’s intrinsic CIRS, anti-inflammatory, and antioxidant capabilities. Overall, there is no evidence that TRYCAT-associated neurotoxicity causes SCZ, while it is not excluded that neurotoxic TRYCATs cause a worsening of FES/MES and hence a deficit SCZ.

**Figure 5:**
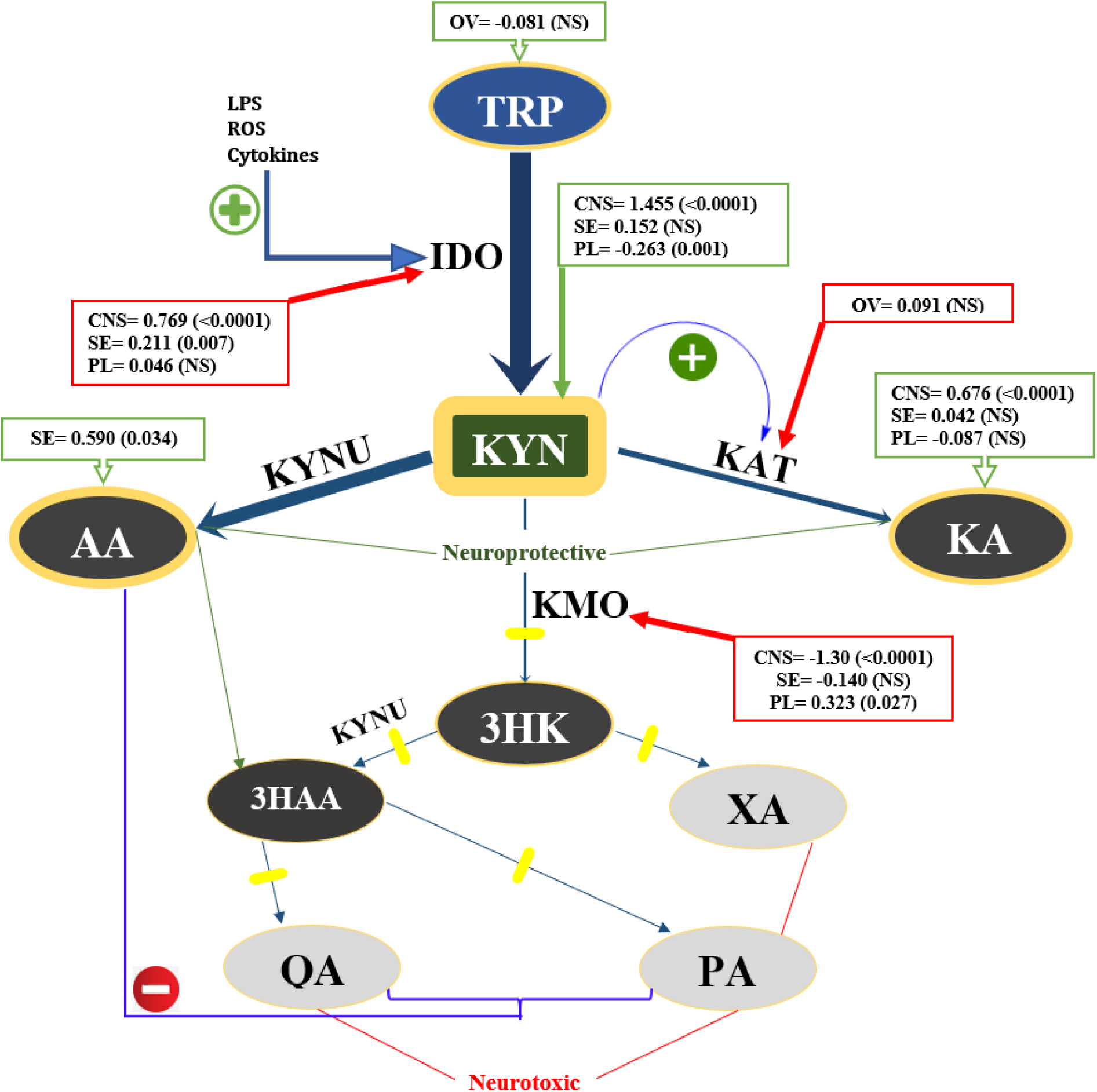
Summary of the findings in schizophrenia. TRP: Tryptophan, KYN: Kynurenine, KA: Kynurenic acid, AA: Anthranilic acid, 3HK: 3-Hydroxy kynurenine, 3HAA: 3-Hydroxy anthranilic acid, XA: Xanthurenic acid, QA: Quinolinic acid, PA: Picolinic acid. IDO: Indoleamine 2,3-dioxygenase, KAT: Kynurenine aminotransferase, KMO: Kynurenine 3-monooxygenase, KYNU: Kynureninase, LPS: Lipopolysaccharides, ROS: Reactive oxygen species. In this graph we also show the effect sizes (as standardized mean difference (SMD) with p-values) for the tryptophan catabolites, namely as OV: Overall, when there were no group differences and as CNS: Central nervous system, SE: Serum, and PL: Plasma, when there were significant group differences. The activities of IDO, KAT and KMO enzymes were estimated by computing ratios, i.e., IDO: KYN/TRP ratio, KAT: (KA+KAT)/KYN ratio, and KMO: (3HK+KMO)/KYN ratio.

## Data Availability

All data produced are available online at PubMed and Google scholar.

## Ethical approval and consent to participate

Not applicable.

## Consent for publication

Not applicable.

## Availability of data and materials

The dataset (excel file) generated during and/or analyzed during the current study will be available from MM upon reasonable request and once the dataset has been fully exploited by the authors.

## Author’s contributions

All authors contributed to the writing up of the paper. The work was designed by AA and MM. Data were collected by AA and AV. Statistical analyses were performed by AA and MM. All authors revised and approved the final draft.

## Acknowledgements

Not applicable.

## Supplementary Information

Supplementary information is available at MP’s website.

## References

1. Smith RS, Maes M. The macrophage-T-lymphocyte theory of schizophrenia: additional evidence. Medical hypotheses 1995; 45(2): 135–141.

2. Kanchanatawan B, Sirivichayakul S, Ruxrungtham K, Carvalho AF, Geffard M, Ormstad H et al. Deficit, but Not Nondeficit, Schizophrenia Is Characterized by Mucosa-Associated Activation of the Tryptophan Catabolite (TRYCAT) Pathway with Highly Specific Increases in IgA Responses Directed to Picolinic, Xanthurenic, and Quinolinic Acid. Mol Neurobiol 2018; 55(2): 1524–1536.

3. Noto C, Maes M, Ota VK, Teixeira AL, Bressan RA, Gadelha A et al. High predictive value of immune-inflammatory biomarkers for schizophrenia diagnosis and association with treatment resistance. World J Biol Psychiatry 2015; 16(6): 422–429.

4. Rubesa G, Gudelj L, Makovac D. Immunological Characteristics of Schizophrenia. Psychiatria Danubina 2018; 30(Suppl 4): 180–187.

5. Al-Hakeim HK, Almulla AF, Maes M. The Neuroimmune and Neurotoxic Fingerprint of Major Neurocognitive Psychosis or Deficit Schizophrenia: a Supervised Machine Learning Study. Neurotox Res 2020; 37(3): 753–771.

6. Almulla AF, Al-Hakeim HK, Abed MS, Carvalho AF, Maes M. Chronic fatigue and fibromyalgia symptoms are key components of deficit schizophrenia and are strongly associated with activated immune-inflammatory pathways. Schizophr Res 2020; 222: 342–353.

7. Maes M, Sirivichayakul S, Matsumoto AK, Maes A, Michelin AP, de Oliveira Semeão L et al. Increased Levels of Plasma Tumor Necrosis Factor-α Mediate Schizophrenia Symptom Dimensions and Neurocognitive Impairments and Are Inversely Associated with Natural IgM Directed to Malondialdehyde and Paraoxonase 1 Activity. Mol Neurobiol 2020; 57(5): 2333–2345.

8. Noto MN, Maes M, Nunes SOV, Ota VK, Rossaneis AC, Verri WA, Jr. et al. Activation of the immune-inflammatory response system and the compensatory immune-regulatory system in antipsychotic naive first episode psychosis. European neuropsychopharmacology : the journal of the European College of Neuropsychopharmacology 2019; 29(3): 416–431.

9. Roomruangwong C, Noto C, Kanchanatawan B, Anderson G, Kubera M, Carvalho AF et al. The Role of Aberrations in the Immune-Inflammatory Response System (IRS) and the Compensatory Immune-Regulatory Reflex System (CIRS) in Different Phenotypes of Schizophrenia: the IRS-CIRS Theory of Schizophrenia. Mol Neurobiol 2020; 57(2): 778–797.

10. Saito K, Markey SP, Heyes MP. Effects of immune activation on quinolinic acid and neuroactive kynurenines in the mouse. Neuroscience 1992; 51(1): 25–39.

11. Reyes Ocampo J, Lugo Huitron R, Gonzalez-Esquivel D, Ugalde-Muniz P, Jimenez-Anguiano A, Pineda B et al. Kynurenines with neuroactive and redox properties: relevance to aging and brain diseases. Oxid Med Cell Longev 2014; 2014: 646909.

12. Anderson G, Maes M. Schizophrenia: linking prenatal infection to cytokines, the tryptophan catabolite (TRYCAT) pathway, NMDA receptor hypofunction, neurodevelopment and neuroprogression. Prog Neuropsychopharmacol Biol Psychiatry 2013; 42: 5–19.

13. Maes M, Fišar Z, Medina M, Scapagnini G, Nowak G, Berk M. New drug targets in depression: inflammatory, cell-mediated immune, oxidative and nitrosative stress, mitochondrial, antioxidant, and neuroprogressive pathways. And new drug candidates— Nrf2 activators and GSK-3 inhibitors. Inflammopharmacology 2012; 20(3): 127–150.

14. Maes M, Leonard BE, Myint AM, Kubera M, Verkerk R. The new ‘5-HT’ hypothesis of depression: Cell-mediated immune activation induces indoleamine 2,3-dioxygenase, which leads to lower plasma tryptophan and an increased synthesis of detrimental tryptophan catabolites (TRYCATs), both of which contribute to the onset of depression. Progress in Neuro-Psychopharmacology and Biological Psychiatry 2011; 35(3): 702–721.

15. Maes M, Mihaylova I, Ruyter MD, Kubera M, Bosmans E. The immune effects of TRYCATs (tryptophan catabolites along the IDO pathway): relevance for depression - and other conditions characterized by tryptophan depletion induced by inflammation. Neuro endocrinology letters 2007; 28(6): 826–831.

16. Krause D, Suh H-S, Tarassishin L, Cui QL, Durafourt BA, Choi N et al. The tryptophan metabolite 3-hydroxyanthranilic acid plays anti-inflammatory and neuroprotective roles during inflammation: role of hemeoxygenase-1. Am J Pathol 2011; 179(3): 1360–1372.

17. Savitz J, Drevets WC, Smith CM, Victor TA, Wurfel BE, Bellgowan PS et al. Putative neuroprotective and neurotoxic kynurenine pathway metabolites are associated with hippocampal and amygdalar volumes in subjects with major depressive disorder. Neuropsychopharmacology 2015; 40(2): 463–471.

18. Baumann P, (ed) Relationships Between Plasma, CSF and Brain Tryptophan. Proceedings of the Transport Mechanisms of Tryptophan in Blood Cells, Nerve Cells, and at the Blood-Brain Barrier; 1979// 1979; Vienna. Springer Vienna.

19. Fernstrom JD, Larin F, Wurtman RJ. Correlation between brain tryptophan and plasma neutral amino acid levels following food consumption in rats. Life Sciences 1973; 13(5): 517–524.

20. Fukui S, Schwarcz R, Rapoport SI, Takada Y, Smith QR. Blood-brain barrier transport of kynurenines: implications for brain synthesis and metabolism. Journal of neurochemistry 1991; 56(6): 2007–2017.

21. Saito K, Fujigaki S, Heyes MP, Shibata K, Takemura M, Fujii H et al. Mechanism of increases in L-kynurenine and quinolinic acid in renal insufficiency. Am J Physiol Renal Physiol 2000; 279(3): F565–572.

22. Topczewska-Bruns J, Pawlak D, Tankiewicz A, Chabielska E, Buczko W. Kynurenine metabolism in central nervous system in experimental chronic renal failure. Adv Exp Med Biol 2003; 527: 177–182.

23. Kita T, Morrison PF, Heyes MP, Markey SP. Effects of systemic and central nervous system localized inflammation on the contributions of metabolic precursors to the L-kynurenine and quinolinic acid pools in brain. Journal of neurochemistry 2002; 82(2): 258–268.

24. Gál EM, Sherman AD. L-kynurenine: its synthesis and possible regulatory function in brain. Neurochemical research 1980; 5(3): 223–239.

25. Linderholm KR, Skogh E, Olsson SK, Dahl ML, Holtze M, Engberg G et al. Increased levels of kynurenine and kynurenic acid in the CSF of patients with schizophrenia. Schizophr Bull 2010; 38(3): 426–432.

26. Schwarcz R, Rassoulpour A, Wu H-Q, Medoff D, Tamminga CA, Roberts RC. Increased cortical kynurenate content in schizophrenia. Biological Psychiatry 2001; 50(7): 521–530.

27. Kindler J, Lim CK, Weickert CS, Boerrigter D, Galletly C, Liu D et al. Dysregulation of kynurenine metabolism is related to proinflammatory cytokines, attention, and prefrontal cortex volume in schizophrenia. Mol Psychiatry 2020; 25(11): 2860–2872.

28. Bonaccorso S, Marino V, Puzella A, Pasquini M, Biondi M, Artini M et al. Increased depressive ratings in patients with hepatitis C receiving interferon-alpha-based immunotherapy are related to interferon-alpha-induced changes in the serotonergic system. J Clin Psychopharmacol 2002; 22(1): 86–90.

29. Lavebratt C, Olsson S, Backlund L, Frisén L, Sellgren C, Priebe L et al. The KMO allele encoding Arg452 is associated with psychotic features in bipolar disorder type 1, and with increased CSF KYNA level and reduced KMO expression. Molecular Psychiatry 2014; 19(3): 334–341.

30. Plitman E, Iwata Y, Caravaggio F, Nakajima S, Chung JK, Gerretsen P et al. Kynurenic Acid in Schizophrenia: A Systematic Review and Meta-analysis. Schizophr Bull 2017; 43(4): 764–777.

31. Morrens M, De Picker L, Kampen JK, Coppens V. Blood-based kynurenine pathway alterations in schizophrenia spectrum disorders: A meta-analysis. Schizophr Res 2020; 223: 43–52.

32. Cao B, Chen Y, Ren Z, Pan Z, McIntyre RS, Wang D. Dysregulation of kynurenine pathway and potential dynamic changes of kynurenine in schizophrenia: A systematic review and meta-analysis. Neurosci Biobehav Rev 2021; 123: 203–214.

33. Marx W, McGuinness AJ, Rocks T, Ruusunen A, Cleminson J, Walker AJ et al. The kynurenine pathway in major depressive disorder, bipolar disorder, and schizophrenia: a meta-analysis of 101 studies. Mol Psychiatry 2021; 26(8): 4158–4178.

34. Page MJ, McKenzie JE, Bossuyt PM, Boutron I, Hoffmann TC, Mulrow CD, et al. The PRISMA 2020 statement: An updated guideline for reporting systematic reviews. PLoS Med 2021; 18(3): e1003583–e1003583.

35. Higgins JPT TJ, Chandler J, Cumpston M, Li T, Page MJ, Welch VA. Cochrane Handbook for Systematic Reviews of Interventions. 2nd Edition edn. John Wiley & Sons: Chichester (UK), 2019.

36. Sathyasaikumar KV, Stachowski EK, Wonodi I, Roberts RC, Rassoulpour A, McMahon RP et al. Impaired kynurenine pathway metabolism in the prefrontal cortex of individuals with schizophrenia. Schizophr Bull 2011; 37(6): 1147–1156.

37. Wan X, Wang W, Liu J, Tong T. Estimating the sample mean and standard deviation from the sample size, median, range and/or interquartile range. BMC Medical Research Methodology 2014; 14(1): 135.

38. Andrés-Rodríguez L, Borràs X, Feliu-Soler A, Pérez-Aranda A, Angarita-Osorio N, Moreno-Peral P et al. Peripheral immune aberrations in fibromyalgia: A systematic review, meta-analysis and meta-regression. Brain, Behavior, and Immunity 2019; 87: 881–889

39. Cohen J. Statistical power analysis for the behavioral sciences. Academic press2013.

40. Vasupanrajit A, Jirakran K, Tunvirachaisakul C, Maes M. Suicide attempts are associated with activated immune-inflammatory, nitro-oxidative, and neurotoxic pathways: A systematic review and meta-analysis. Journal of Affective Disorders 2021; 295: 80–92.

41. Vasupanrajit A, Jirakran K, Tunvirachaisakul C, Solmi M, Maes M. Inflammation and nitro-oxidative stress in current suicidal attempts and current suicidal ideation: a systematic review and meta-analysis. medRxiv 2021: 2021.2009.2009.21263363.

42. Wurfel BE, Drevets WC, Bliss SA, McMillin JR, Suzuki H, Ford BN et al. Serum kynurenic acid is reduced in affective psychosis. Transl Psychiatry 2017; 7(5): e1115.

43. Szymona K, Zdzisinska B, Karakula-Juchnowicz H, Kocki T, Kandefer-Szerszen M, Flis M et al. Correlations of Kynurenic Acid, 3-Hydroxykynurenine, sIL-2R, IFN-alpha, and IL-4 with Clinical Symptoms During Acute Relapse of Schizophrenia. Neurotox Res 2017; 32(1): 17–26.

44. Schwieler L, Larsson MK, Skogh E, Kegel ME, Orhan F, Abdelmoaty S et al. Increased levels of IL-6 in the cerebrospinal fluid of patients with chronic schizophrenia--significance for activation of the kynurenine pathway. J Psychiatry Neurosci 2015; 40(2): 126–133.

45. Nilsson LK, Linderholm KR, Engberg G, Paulson L, Blennow K, Lindstrom LH et al. Elevated levels of kynurenic acid in the cerebrospinal fluid of male patients with schizophrenia. Schizophr Res 2005; 80(2-3): 315–322.

46. Myint AM, Schwarz MJ, Verkerk R, Mueller HH, Zach J, Scharpe S et al. Reversal of imbalance between kynurenic acid and 3-hydroxykynurenine by antipsychotics in medication-naive and medication-free schizophrenic patients. Brain Behav Immun 2011; 25(8): 1576–1581.

47. Huang X, Ding W, Wu F, Zhou S, Deng S, Ning Y. Increased Plasma Kynurenic Acid Levels are Associated with Impaired Attention/Vigilance and Social Cognition in Patients with Schizophrenia. Neuropsychiatr Dis Treat 2020; 16: 263–271.

48. Huang J, Tong J, Zhang P, Zhou Y, Cui Y, Tan S et al. Effects of neuroactive metabolites of the tryptophan pathway on working memory and cortical thickness in schizophrenia. Transl Psychiatry 2021; 11(1): 198.

49. Holtze M, Saetre P, Engberg G, Schwieler L, Werge T, Andreassen OA et al. Kynurenine 3-monooxygenase polymorphisms: relevance for kynurenic acid synthesis in patients with schizophrenia and healthy controls. J Psychiatry Neurosci 2012; 37(1): 53–57.

50. Erhardt S, Blennow K, Nordin C, Skogh E, Lindström LH, Engberg G. Kynurenic acid levels are elevated in the cerebrospinal fluid of patients with schizophrenia. Neuroscience Letters 2001; 313(1-2): 96–98.

51. De Picker L, Fransen E, Coppens V, Timmers M, de Boer P, Oberacher H et al. Immune and Neuroendocrine Trait and State Markers in Psychotic Illness: Decreased Kynurenines Marking Psychotic Exacerbations. Front Immunol 2019; 10: 2971.

52. Chiappelli J, Postolache TT, Kochunov P, Rowland LM, Wijtenburg SA, Shukla DK et al. Tryptophan Metabolism and White Matter Integrity in Schizophrenia. Neuropsychopharmacology 2016; 41(10): 2587–2595.

53. Chiappelli J, Notarangelo FM, Pocivavsek A, Thomas MAR, Rowland LM, Schwarcz R et al. Influence of plasma cytokines on kynurenine and kynurenic acid in schizophrenia. Neuropsychopharmacology 2018; 43(8): 1675–1680.

54. Cathomas F, Guetter K, Seifritz E, Klaus F, Kaiser S. Quinolinic acid is associated with cognitive deficits in schizophrenia but not major depressive disorder. Sci Rep 2021; 11(1): 9992.

55. Barry S, Clarke G, Scully P, Dinan TG. Kynurenine pathway in psychosis: evidence of increased tryptophan degradation. J Psychopharmacol 2009; 23(3): 287–294.

56. Cao B, Wang D, Brietzke E, McIntyre RS, Pan Z, Cha D et al. Characterizing amino-acid biosignatures amongst individuals with schizophrenia: a case-control study. Amino Acids 2018; 50(8): 1013–1023.

57. Carl GF, Brogan MP, Young BK. Is plasma serine a marker for psychosis? Biological Psychiatry 1992; 31(11): 1130–1135.

58. Condray R, Dougherty GG, Jr., Keshavan MS, Reddy RD, Haas GL, Montrose DM et al. 3-Hydroxykynurenine and clinical symptoms in first-episode neuroleptic-naive patients with schizophrenia. Int J Neuropsychopharmacol 2011; 14(6): 756–767.

59. Curto M, Lionetto L, Fazio F, Corigliano V, Comparelli A, Ferracuti S et al. Serum xanthurenic acid levels: Reduced in subjects at ultra high risk for psychosis. Schizophr Res 2019; 208: 465–466.

60. Fazio F, Lionetto L, Curto M, Iacovelli L, Cavallari M, Zappulla C et al. Xanthurenic Acid Activates mGlu2/3 Metabotropic Glutamate Receptors and is a Potential Trait Marker for Schizophrenia. Sci Rep 2015; 5: 17799.

61. Fukushima T, Iizuka H, Yokota A, Suzuki T, Ohno C, Kono Y et al. Quantitative analyses of schizophrenia-associated metabolites in serum: serum D-lactate levels are negatively correlated with gamma-glutamylcysteine in medicated schizophrenia patients. PLoS One 2014; 9(7): e101652.

62. Joaquim HPG, Costa AC, Gattaz WF, Talib LL. Kynurenine is correlated with IL-1beta in plasma of schizophrenia patients. J Neural Transm (Vienna) 2018; 125(5): 869–873.

63. Kegel ME, Bhat M, Skogh E, Samuelsson M, Lundberg K, Dahl ML et al. Imbalanced kynurenine pathway in schizophrenia. Int J Tryptophan Res 2014; 7: 15–22.

64. Kim YK, Myint AM, Verkerk R, Scharpe S, Steinbusch H, Leonard B. Cytokine changes and tryptophan metabolites in medication-naive and medication-free schizophrenic patients. Neuropsychobiology 2009; 59(2): 123–129.

65. Lee M, Jayathilake K, Dai J, Meltzer HY. Decreased plasma tryptophan and tryptophan/large neutral amino acid ratio in patients with neuroleptic-resistant schizophrenia: relationship to plasma cortisol concentration. Psychiatry Res 2011; 185(3): 328–333.

66. Leppik L, Kriisa K, Koido K, Koch K, Kajalaid K, Haring L et al. Profiling of Amino Acids and Their Derivatives Biogenic Amines Before and After Antipsychotic Treatment in First-Episode Psychosis. Front Psychiatry 2018; 9: 155.

67. Oxenkrug G, van der Hart M, Roeser J, Summergrad P. Peripheral kynurenine-3-monooxygenase deficiency as a potential risk factor for metabolic syndrome in schizophrenia patients. Integr Clin Med 2017; 1(1).

68. Paredes RM, Quinones M, Marballi K, Gao X, Valdez C, Ahuja SS et al. Metabolomic profiling of schizophrenia patients at risk for metabolic syndrome. Int J Neuropsychopharmacol 2014; 17(8): 1139–1148.

69. Rao ML, Gross G, Strebel B, Bräunig P, Huber G, Klosterkötter J. Serum amino acids, central monoamines, and hormones in drug-naive, drug-free, and neuroleptic-treated schizophrenic patients and healthy subjects. Psychiatry Research 1990; 34(3): 243–257.

70. Ravikumar A, Deepadevi KV, Arun P, Manojkumar V, Kurup PA. Tryptophan and tyrosine catabolic pattern in neuropsychiatric disorders. Neurol India 2000; 48(3): 231–238.

71. Tortorella A, Monteleone P, Fabrazzo M, Viggiano A, De Luca L, Maj M. Plasma concentrations of amino acids in chronic schizophrenics treated with clozapine. Neuropsychobiology 2001; 44(4): 167–171.

72. van der Heijden FM, Fekkes D, Tuinier S, Sijben AE, Kahn RS, Verhoeven WM. Amino acids in schizophrenia: evidence for lower tryptophan availability during treatment with atypical antipsychotics? J Neural Transm (Vienna) 2005; 112(4): 577–585.

73. Zhang Z, Zhang M, Luo Y, Ni X, Lu H, Wen Y et al. Preliminary comparative analysis of kynurenine pathway metabolites in chronic ketamine users, schizophrenic patients, and healthy controls. Hum Psychopharmacol 2020; 35(4): e2738.

74. Yao JK, Dougherty GG, Jr., Reddy RD, Keshavan MS, Montrose DM, Matson WR et al. Altered interactions of tryptophan metabolites in first-episode neuroleptic-naive patients with schizophrenia. Mol Psychiatry 2010; 15(9): 938–953.

75. Wu F, Li H, Zhou Y, Huang Y. Relationship between kynurenine pathway metabolites and cognitive function in schizophrenia patients. Clin Med Eng 2019; 26: 1153–1154.

76. Wei J. Low concentrations of serum tyrosine in neuroleptic-free schizophrenics with an early onset. Schizophrenia Research 1995; 14(3): 257–260.

77. van de Kerkhof NW, Fekkes D, van der Heijden FM, Hoogendijk WJ, Stober G, Egger JI et al. Cycloid psychoses in the psychosis spectrum: evidence for biochemical differences with schizophrenia. Neuropsychiatr Dis Treat 2016; 12: 1927–1933.

78. Tang Y, Chen T, Zhang X, Luo X, Liu Y. Determination and clinical significance of serum kynurenine and kynurenic acid levels in schizophrenia %. J Chin J Behav Med Sci 2009: 103–104.

79. Steen NE, Dieset I, Hope S, Vedal TSJ, Smeland OB, Matson W et al. Metabolic dysfunctions in the kynurenine pathway, noradrenergic and purine metabolism in schizophrenia and bipolar disorders. Psychol Med 2020; 50(4): 595–606.

80. Sperner-Unterweger B, Miller C, Holzner B, Laich A, Widner B, Fleischhacker WW, et al. Immunologic Alterations in Schizophrenia: Neopterin, L-Kynurenine, Tryptophan and T-Cell Subsets in the Acute Stage of Illness %J Pteridines. 2002; 13(1): 9–14.

81. Rao ML, Gross G, Strebel B, Halaris A, Huber G, Bräunig P et al. Circadian rhythm of tryptophan, serotonin, melatonin, and pituitary hormones in schizophrenia. Biol Psychiatry 1994; 35(3): 151–163.

82. Oxenkrug G, van der Hart M, Roeser J, Summergrad P. Anthranilic Acid: A Potential Biomarker and Treatment Target for Schizophrenia. Ann Psychiatry Ment Health 2016; 4(2): 1059.

83. Noyan H, Erdag E, Tuzun E, Yaylim I, Kucukhuseyin O, Hakan MT et al. Association of the kynurenine pathway metabolites with clinical, cognitive features and IL-1beta levels in patients with schizophrenia spectrum disorder and their siblings. Schizophr Res 2021; 229: 27–37.

84. Nilsson-Todd LK, Nordin C, Jonsson EG, Skogh E, Erhardt S. Cerebrospinal fluid kynurenic acid in male patients with schizophrenia - correlation with monoamine metabolites. Acta Neuropsychiatr 2007; 19(1): 45–52.

85. Manowitz P, Gilmour DG, Racevskis J. Low plasma tryptophan levels in recently hospitalized schizophrenics. Biological Psychiatry 1973; 6(2): 109–118.

86. Koike S, Bundo M, Iwamoto K, Suga M, Kuwabara H, Ohashi Y et al. A snapshot of plasma metabolites in first-episode schizophrenia: a capillary electrophoresis time-of-flight mass spectrometry study. Translational Psychiatry 2014; 4(4): e379–e379.

87. Glassman M, Wehring HJ, Pocivavsek A, Sullivan KM, Rowland LM, McMahon RP et al. Peripheral Cortisol and Inflammatory Response to a Psychosocial Stressor in People with Schizophrenia. J Neuropsychiatry (Foster City) 2018; 2(2).

88. Domino EF, Krause RR. Plasma tryptophan tolerance curves in drug free normal controls, schizophrenic patients and prisoner volunteers. Journal of psychiatric research 1974; 10(3-4): 247–261.

89. Brundin L, Sellgren CM, Lim CK, Grit J, Palsson E, Landen M et al. An enzyme in the kynurenine pathway that governs vulnerability to suicidal behavior by regulating excitotoxicity and neuroinflammation. Transl Psychiatry 2016; 6(8): e865.

90. Oxenkrug G, Bernstein HG, Guest PC, van der Hart M, Roeser J, Summergrad P et al. Plasma xanthurenic acid in a context of insulin resistance and obesity in schizophrenia. Schizophr Res 2019; 211: 98–99.

91. Potkin SG, Cannon-Spoor HE, DeLisi LE, Neckers LM, Wyatt RJ. Plasma phenylalanine, tyrosine, and tryptophan in schizophrenia. Arch Gen Psychiatry 1983; 40(7): 749–752.

92. Fekkes D, Bode WT, Zijlstra FJ, Pepplinkhuizen L. Eicosanoid and amino acid metabolism in transient acute psychoses with psychedelic symptoms. Prostaglandins, leukotrienes, and essential fatty acids 1996; 54(4): 261–264.

93. Miller CL, Llenos IC, Dulay JR, Weis S. Upregulation of the initiating step of the kynurenine pathway in postmortem anterior cingulate cortex from individuals with schizophrenia and bipolar disorder. Brain Res 2006; 1073-1074: 25-37.

94. Pi G, Tang A, Mo X, Luo X, Ao X. Determination of kynurenic acid and tryptophan in serum by high performance liquid chromatography with fluorescence detection %. J Chin J Lab Med 2007; 30: 1134–1137.

95. Domingues DS, Crevelin EJ, de Moraes LA, Cecilio Hallak JE, de Souza Crippa JA, Costa Queiroz ME. Simultaneous determination of amino acids and neurotransmitters in plasma samples from schizophrenic patients by hydrophilic interaction liquid chromatography with tandem mass spectrometry. J Sep Sci 2015; 38(5): 780–787.

96. Yang X, Yang Y, Lang W, Liu Y, Chen Z, Tao H. Determination and clinical significance of serum kynurenine and kynurenine 3 -monooxygenase levels in schizophrenia. J Hunan Normal Univ (Med Sci) 2016; 13: 18–19.

97. Shovestul BJ, Glassman M, Rowland LM, McMahon RP, Liu F, Kelly DL. Pilot study examining the relationship of childhood trauma, perceived stress, and medication use to serum kynurenic acid and kynurenine levels in schizophrenia. Schizophr Res 2017; 185: 200–201.

98. Okamoto N, Natsuyama T, Igata R, Konishi Y, Tesen H, Ikenouchi A et al. Associations Between the Kynurenine Pathway, Proinflammatory Cytokines, and Brain-Derived Neurotrophic Factor in Hospitalized Patients With Chronic Schizophrenia: A Preliminary Study. Front Psychiatry 2021; 12: 696059.

99. Anderson G, Maes M. How Immune-inflammatory Processes Link CNS and Psychiatric Disorders: Classification and Treatment Implications. CNS & Neurological Disorders - Drug Targets- CNS & Neurological Disorders) 2017; 16(3): 266–278.

100. Anderson G, Maes M. Interactions of Tryptophan and Its Catabolites With Melatonin and the Alpha 7 Nicotinic Receptor in Central Nervous System and Psychiatric Disorders: Role of the Aryl Hydrocarbon Receptor and Direct Mitochondria Regulation. Int J Tryptophan Res 2017; 10: 1178646917691738.

101. Kanchanatawan B, Sirivichayakul S, Ruxrungtham K, Carvalho AF, Geffard M, Ormstad H et al. Deficit, but Not Nondeficit, Schizophrenia Is Characterized by Mucosa-Associated Activation of the Tryptophan Catabolite (TRYCAT) Pathway with Highly Specific Increases in IgA Responses Directed to Picolinic, Xanthurenic, and Quinolinic Acid. Molecular Neurobiology 2018; 55(2): 1524–1536.

102. Maes M, Galecki P, Verkerk R, Rief W. Somatization, but not depression, is characterized by disorders in the tryptophan catabolite (TRYCAT) pathway, indicating increased indoleamine 2,3-dioxygenase and lowered kynurenine aminotransferase activity. Neuroendocrinology Letters 2011; 32(3): 264–273.

103. Nantachai G, Vasupanrajit A, Tunvirachaisakul C, Solmi M, Maes M. Oxidative stress and antioxidant defenses in mild cognitive impairment: a systematic review and meta-analysis. 2021: 2021.2011.2022.21266698.

104. Schwarcz R, Bruno JP, Muchowski PJ, Wu HQ. Kynurenines in the mammalian brain: when physiology meets pathology. Nature reviews Neuroscience 2012; 13(7): 465–477.

105. Reyes Ocampo J, Lugo Huitrón R, González-Esquivel D, Ugalde-Muñiz P, Jiménez-Anguiano A, Pineda B et al. Kynurenines with Neuroactive and Redox Properties: Relevance to Aging and Brain Diseases. Oxidative Medicine and Cellular Longevity 2014; 2014: 646909.

106. Guillemin GJ, Kerr SJ, Smythe GA, Smith DG, Kapoor V, Armati PJ et al. Kynurenine pathway metabolism in human astrocytes: a paradox for neuronal protection. Journal of neurochemistry 2001; 78(4): 842–853.

107. Speciale C, Wu H-Q, Cini M, Marconi M, Varasi M, Schwarcz RJEjop. (R, S)-3, 4-dichlorobenzoylalanine (FCE 28833A) causes a large and persistent increase in brain kynurenic acid levels in rats. 1996; 315(3): 263–267.

108. Röver S, Cesura AM, Huguenin P, Kettler R, Szente AJJomc. Synthesis and biochemical evaluation of N-(4-phenylthiazol-2-yl) benzenesulfonamides as high-affinity inhibitors of kynurenine 3-hydroxylase. 1997; 40(26): 4378–4385.

109. Darlington LG, Forrest CM, Mackay GM, Smith RA, Smith AJ, Stoy N et al. On the Biological Importance of the 3-hydroxyanthranilic Acid: Anthranilic Acid Ratio. Int J Tryptophan Res 2010; IJTR 3: 51–59.

110. Gregory Oxenkrug MvdH, Julien Roeser, and Paul Summergrad. Anthranilic Acid: A Potential Biomarker and Treatment Target for Schizophrenia. 2016; 4(2).

111. Guillemin GJ, Cullen KM, Lim CK, Smythe GA, Garner B, Kapoor V et al. Characterization of the kynurenine pathway in human neurons. The Journal of neuroscience : the official journal of the Society for Neuroscience 2007; 27(47): 12884–12892.

112. Wirthgen E, Hoeflich A, Rebl A, Gunther J. Kynurenic Acid: The Janus-Faced Role of an Immunomodulatory Tryptophan Metabolite and Its Link to Pathological Conditions. Front Immunol 2017; 8: 1957.

113. Davies NWS, Guillemin G, Brew BJ. Tryptophan, Neurodegeneration and HIV-Associated Neurocognitive Disorder. 2010; 3: IJTR.S4321.

114. Banerjee J, Alkondon M, Pereira EFR, Albuquerque EX. Regulation of GABAergic inputs to CA1 pyramidal neurons by nicotinic receptors and kynurenic acid. J Pharmacol Exp Ther 2012; 341(2): 500–509.

115. Maes M, Vojdani A, Sirivichayakul S, Barbosa DS, Kanchanatawan B. Inflammatory and Oxidative Pathways Are New Drug Targets in Multiple Episode Schizophrenia and Leaky Gut, Klebsiella pneumoniae, and C1q Immune Complexes Are Additional Drug Targets in First Episode Schizophrenia. Mol Neurobiol 2021; 58(7): 3319–3334.

116. Sotelo-Orozco J, Chen S-Y, Hertz-Picciotto I, Slupsky CM. A Comparison of Serum and Plasma Blood Collection Tubes for the Integration of Epidemiological and Metabolomics Data. 2021; 8(650).

117. Parvy PR, Bardet JI, Kamoun PP. EDTA in vacutainer tubes can interfere with plasma amino acid analysis. Clinical chemistry 1983; 29(4): 735.

118. Bellmaine S, Schnellbaecher A, Zimmer A. Reactivity and degradation products of tryptophan in solution and proteins. Free Radical Biology and Medicine 2020; 160: 696–718.

119. Kulkarni P, Karanam A, Gurjar M, Dhoble S, Naik AB, Vidhun BH et al. Effect of various anticoagulants on the bioanalysis of drugs in rat blood: implication for pharmacokinetic studies of anticancer drugs. Springerplus 2016; 5(1): 2102–2102.

120. Davidson DF. Effects of contamination of blood specimens with liquid potassium-EDTA anticoagulant. Annals of clinical biochemistry 2002; 39(Pt 3): 273–280.

121. Baumann P, Perey M. The analysis of free tryptophan in human blood with the Ultrafiltrator: a comparison with other methods. Clinica chimica acta; international journal of clinical chemistry 1977; 76(2): 223–231.

122. Bourgoin B, Faivre-Bauman A, Hery F, Ternaux JP, Hamon M. Characteristics of tryptophan binding in the serum of the newborn rat. Biology of the neonate 1977; 31(3-4): 141–154.

123. Dietrich-Muszalska A, Kwiatkowska A. Generation of superoxide anion radicals and platelet glutathione peroxidase activity in patients with schizophrenia. Neuropsychiatr Dis Treat 2014; 10: 703–709.

124. Quintana J. Platelet serotonin and plasma tryptophan decreases in endogenous depression. Clinical, therapeutic, and biological correlations. Journal of Affective Disorders 1992; 24(2): 55–62.

125. McMenamy RH, Watson F. Indole-albumin association: A comparative study. Comparative Biochemistry and Physiology 1968; 26(1): 329–335.

126. Pardridge WM, Fierer G. Transport of tryptophan into brain from the circulating, albumin-bound pool in rats and in rabbits. Journal of neurochemistry 1990; 54(3): 971–976.

127. Maes M, Jacobs MP, Suy E, Vandewoude M, Minner B, Raus J. Effects of dexamethasone on the availability of L-tryptophan and on the insulin and FFA concentrations in unipolar depressed patients. Biol Psychiatry 1990; 27(8): 854–862.

128. Uwai Y, Honjo H, Iwamoto K. Interaction and transport of kynurenic acid via human organic anion transporters hOAT1 and hOAT3. Pharmacol Res 2012; 65(2): 254–260.

