## supplementary file for "The tryptophan catabolite or kynurenine pathway in schizophrenia: meta-analysis reveals dissociations between central, serum and plasma compartments"

### ESF, Table 1. Specific search for each database

| Database Name | Search Sentence | No. of Articles |
|---|---|---|
| PubMed/Medline | ((((((((((((Schizophrenia and TRYCATs)* OR (Schizophrenia and kynurenine)) OR (Schizophrenia and kynurenic acid)) OR (schizophrenia and quinolinic acid)) OR (Schizophrenia and Picolinic acid)) OR (Schizophrenia and 3-hydroxyanthranilic acid)) OR (Schizophrenia and xanthurenic acid)) OR (Schizophrenia and 3-hydroxykynurenine)) OR (Schizophrenia and Anthranilic acid)) OR (Schizophrenia and formyl kynurenine)) OR (Schizophrenia and l-tryptophan)) OR (Schizophrenia and tryptophan catabolites)) OR (TRYCATs and SCZ)) OR (TRYCATs and Psychosis) | **838** |
| | (((((((((((TRYCATs and SCZ) OR (TRYCATs and Psychotic spectrum)) OR (Tryptophan Catabolites and SCZ)) OR (L-TRP and SCZ)) OR (L-KYN and SCZ)) OR (KYNA and SCZ)) OR (ANA and SCZ)) OR (3-HK and SCZ)) OR (XA and SCZ)) OR (3-HANA and SCZ)) OR (QA and SCZ)) OR (PA and SCZ) | **309** |
| | ((((((((((SSD and TRYCATs)* OR (SSD and Tryptophan)) OR (Schizophrenia spectrum and tryptophan cata)) OR (PS and TRYCATs)) OR (Psychotic spectrum and TRP)) OR (SSD and KYN)) OR (Psychotic spectrum and kynurenine)) OR (SSD and Kynurenine pathway)) OR (SCZ and kynurenine pathway)) OR (Schizophrenia and IDO)) OR (Schizophrenia and TDO) | **22** |
| Google Scholar | (((((Schizophrenia* and IDO activation) OR (TDO activation)) OR (Kynurenine pathway)) OR (KMO activation)) OR (Tryptophan degradation)) AND (Decreased Tryptophan) | **2060** |
| | SCZ* AND [TRYCATs*] OR [Tryptophan catabolites*] AND [KYN] AND KYNA OR [Picolinic acid] OR [quinolinic acid] OR [3-hydroxyanthranilic acid] OR [xanthurenic acid] OR [3-hydroxykynurenine] OR [Anthranilic acid] OR [IDO] OR [TDO] OR [KMO] | **3** |
| | Schizophrenia* AND [TRYCATs*] OR [Tryptophan catabolites*] AND [kynurenine] AND kynurenic acid OR [Picolinic acid] OR [quinolinic acid] OR [3-hydroxyanthranilic acid] OR [xanthurenic acid] OR [3-hydroxykynurenine] OR [Anthranilic acid] | **41** |

| | | |
|---|---|---|
| **Web of Science** | (((((((ALL=(schizophrenia, TRYCATs)) OR ALL=(Schizophrenia, Tryptophan catabolites )) OR ALL=(schizophr, kynurenine pathway )) OR ALL=(schizophr, IDO and TDO activity )) OR ALL=(schizophr, xanthurenic acid, picolinic acid))) OR ALL=(schizophr, kynurenic acid and kynurenine )) OR ALL=(schizophr, anthranilic acid and 3-hydroxyanthranlinic acid ) | **70** |
| **Web of Science** | ((TS=(Schizophrenia)) AND ALL=("TRYCATs*" OR "tryptophan catabolites*")) AND ALL=("Kynurenine" OR "KYN" OR "kynurenic acid" OR "3-HK" OR "anthranilic acid" OR "picolinic acid" OR "quinolinic acid" OR "xanthurenic acid" OR "IDO" OR "TDO" OR "KMO" OR "3-Hydroxyanthranilic acid" OR "Kynurenine pathway" OR "Tryptophan") | **37** |

**ESF, Table 2. Immune cofounder's scale (ICS) applied from Andrés-Rodríguez, et al., 2019**

| | |
|---|---|
| **Methodological quality of the study** | |
| **1** | Study sample ≥ 128 participants including patients and controls (1= Yes, 0 = No) |
| **2** | Did the study control the results for potential confounders (e.g., age, BMI, gender, race)? (1= Yes, 0 = No) |
| **3** | Were participants with schizophrenia and controls age- and-gender-matched or was there a statistical control? (1= Yes, 0 = No) |
| **4** | Was the time of sample collection specified (e.g., morning vs. evening)? (1= Yes, 0 = No) |
| **5** | Were participants with schizophrenia free of immunomodulatory drugs including anti-cytokines, glucocorticoids, immunoglobulins, and immunosuppressants, or was there a medication washout period or was drug intake statistically controlled for? (1= Yes, 0 = No) |
| **6** | Were participants with schizophrenia free of antidepressants and mood stabilizers or were the data statistically controlled for? (1= Yes, 0 = No) |
| **7** | Reporting either the manufacturer of the test or detection limit and coefficients of variation (1= Yes, 0 = No) |
| **8** | Reporting how data under detection limit were handled (1 = Yes, 0 = No) |
| **9** | Reporting % of the sample under detection limit (1=Yes, 0= No) |
| **10** | Reporting blood fraction (serum, plasma, culture supernatant or whole blood) (1= Yes, 0 = No) |
| | **Total quality score (10 points)** |
| **Biomarker confounders red points**<br>*The red points should not be given if the item is statistically controlled for* | |
| **1** | 3 red points for comorbid illnesses such as autoimmune disorders & other immune disorders including rheumatoid arthritis, psoriasis, inflammatory bowel disease, chronic obstructive pulmonary disease, multiple sclerosis |
| **2** | 3 red points for use of recreational drugs such as methamphetamine or opioids |
| **3** | 2 red points when groups were not matched for age |
| **4** | 2 red points when groups were not matched for sex |
| **5** | 2 red points for medication use as for example immunomodulators |
| **6** | 2 red points for early traumatic life events |

| | |
|---|---|
| **7** | 2 red points for shift work and primary sleep disorders |
| **8** | 1.5 red points for use of neuroleptics |
| **9** | 1 red point for more common systemic immune disorders including diabetes type 1/2, essential hypertension, metabolic syndrome |
| **10** | 1 red point for not fasting (8 hours before blood extraction) |
| **11** | 1 red point for use of omega-3 and antioxidant supplements |
| **12** | 1 red point when data were not controlled for body mass index |
| **13** | 1 red point when data were not controlled for physical activity or sedentary life |
| **14** | 1 red point when data were not controlled for smoking |
| **15** | 1 red point for use of oral contraceptives or NSAIDs |
| **16** | 0.5 red points when data were not controlled for ethnicity in countries such as US, Brazil |
| **17** | 0.5 red points when data were not controlled for seasonality |
| **18** | 0.5 red points when data were not controlled for diurnal variation (8-10 a.m. versus all other time points) |
| | **Total red point score (26 points)** |

Threshold of study samples is established as it is the minimum needed for statistical power. Confounders red points should be given when the item is not reported (or statistically controlled).
The original scales were reported by Andrés-Rodríguez et al.

**Reference:**

Andrés-Rodríguez, L., Borràs, X., Feliu-Soler, A., Pérez-Aranda, A., Angarita-Osorio, N., Moreno-Peral, P., Montero-Marin, J., García-Campayo, J., Carvalho, A. F., Maes, M., Luciano, J. V., 2019. Peripheral immune aberrations in fibromyalgia: A systematic review, meta-analysis and meta-regression. Brain Behav. Immun. 87, 881–889. https://doi.org/10.1016/j.bbi.2019.12.020.

### ESF, Table 3. PRISMA checklist

| Section/topic | # | Checklist item | Reported on page # |
|---|---|---|---|
| **TITLE** | | | |
| Title | 1 | Identify the report as a systematic review, meta-analysis, or both. | 1 |
| **ABSTRACT** | | | |
| Structured summary | 2 | Provide a structured summary including, as applicable: background; objectives; data sources; study eligibility criteria, participants, and interventions; study appraisal and synthesis methods; results; limitations; conclusions and implications of key findings; systematic review registration number. | 3 |
| **INTRODUCTION** | | | |
| Rationale | 3 | Describe the rationale for the review in the context of what is already known. | 5 |
| Objectives | 4 | Provide an explicit statement of questions being addressed with reference to participants, interventions, comparisons, outcomes, and study design (PICOS). | 7 |
| **METHODS** | | | |
| Protocol and registration | 5 | Indicate if a review protocol exists, if and where it can be accessed (e.g., Web address), and, if available, provide registration information including registration number. | 8 |
| Eligibility criteria | 6 | Specify study characteristics (e.g., PICOS, length of follow-up) and report characteristics (e.g., years considered, language, publication status) used as criteria for eligibility, giving rationale. | 9 |
| Information sources | 7 | Describe all information sources (e.g., databases with dates of coverage, contact with study authors to identify additional studies) in the search and date last searched. | 8 |
| Search | 8 | Present full electronic search strategy for at least one database, including any limits used, such that it could be repeated. | 8 |
| Study selection | 9 | State the process for selecting studies (i.e., screening, eligibility, included in systematic review, and, if applicable, included in the meta-analysis). | 8 |
| Data collection process | 10 | Describe method of data extraction from reports (e.g., piloted forms, independently, in duplicate) and any processes for obtaining and confirming data from investigators. | 10 |
| Data items | 11 | List and define all variables for which data were sought (e.g., PICOS, funding sources) and any assumptions and simplifications made. | 11 |
| Risk of bias in individual studies | 12 | Describe methods used for assessing risk of bias of individual studies (including specification of whether this was done at the study or outcome level), and how this information is to be used in any data synthesis. | 12 |
| Summary measures | 13 | State the principal summary measures (e.g., risk ratio, difference in means). | 12 |

| | | | |
|---|---|---|---|
| Synthesis of results | 14 | Describe the methods of handling data and combining results of studies, if done, including measures of consistency (e.g., $I^2$) for each meta-analysis. | 12 |
| Risk of bias across studies | 15 | Specify any assessment of risk of bias that may affect the cumulative evidence (e.g., publication bias, selective reporting within studies). | 12 |
| Additional analyses | 16 | Describe methods of additional analyses (e.g., sensitivity or subgroup analyses, meta-regression), if done, indicating which were pre-specified. | 12 |
| **RESULTS** | | | |
| Study selection | 17 | Give numbers of studies screened, assessed for eligibility, and included in the review, with reasons for exclusions at each stage, ideally with a flow diagram. | 12 |
| Study characteristics | 18 | For each study, present characteristics for which data were extracted (e.g., study size, PICOS, follow-up period) and provide the citations. | 12 |
| Risk of bias within studies | 19 | Present data on risk of bias of each study and, if available, any outcome level assessment (see item 12). | ESF, 14, table 5 |
| Results of individual studies | 20 | For all outcomes considered (benefits or harms), present, for each study: (a) simple summary data for each intervention group (b) effect estimates and confidence intervals, ideally with a forest plot. | ESF, 8, table 4 |
| Synthesis of results | 21 | Present results of each meta-analysis done, including confidence intervals and measures of consistency. | 41, table 2 |
| Risk of bias across studies | 22 | Present results of any assessment of risk of bias across studies (see Item 15). | ESF, 14, table 5 |
| Additional analysis | 23 | Give results of additional analyses, if done (e.g., sensitivity or subgroup analyses, meta-regression [see Item 16]). | ESF, 16, table 6 |
| **DISCUSSION** | | | |
| Summary of evidence | 24 | Summarize the main findings including the strength of evidence for each main outcome; consider their relevance to key groups (e.g., healthcare providers, users, and policy makers). | 18-24 |
| Limitations | 25 | Discuss limitations at study and outcome level (e.g., risk of bias), and at review-level (e.g., incomplete retrieval of identified research, reporting bias). | 24 |
| Conclusions | 26 | Provide a general interpretation of the results in the context of other evidence, and implications for future research. | 25, Figure 6 |
| **FUNDING** | | | |
| Funding | 27 | Describe sources of funding for the systematic review and other support (e.g., supply of data); role of funders for the systematic review. | 26 |

**ESF, Table 4.** Studies excluded from the meta-analysis but included in the systematic review.

| Authors, year | Reason why excluded from the meta-analysis | Results |
|---|---|---|
| Kanchanatawan et al. 2018 | This study exmanied the immune IgA response aganist TRYCATs in schizophrenia | IgA to neurotoxic tryptophan catabolites (TRYCATs) (picolinic, quinolinic, and xanthurenic acid) are significantly increased in deficit schizophrenia. |
| Kanchanatawan et al. 2019 | This study exmanied the immune (IgA/IgM) response aganist TRYCATs in schizophrenia | The results showed increased IgA but lowered IgM against TRYCATs in deficit schizophrenia. |
| Kegel 2017 | No control group. | There is an association between CSF kynureniv acid (KA) levels and psychotic symptoms and personality traits, strengthening the role of KA as a pathophysiological component in psychotic symptomatology |
| Chiappelli et al. 2014 | The study utilized saliva for the assay of KA (exclusion criterion) | Schizophrenia patients show higher levels of salivary KA. |
| Chiappelli et al. 2018 | The study utilized saliva for assay KA | The results indicates a relationship of KA with central glutamatergic dysfunction in response to stress. |
| Aymard et al. 1999 | The study assayed tryptophan (TRP) in whole blood (exclusion criterion) | No significant difference in TRP levels |
| Freedman et al. 1981 | The study recruit a whole blood for determine TRP, also within our exclusion criteria | The results indicate there is a significant decrease in TRP in schiphrenia |

**ESF, Table 5.** Studies included in the meta-analysis

| NO. | Authors, years | Setting | Type of case | Type of Control | N | | | Age | | Assessed Biomarkers | Specimen | Method | Episode | Med | Quality score | Red point score | Results |
|---|---|---|---|---|---|---|---|---|---|---|---|---|---|---|---|---|---|
| | | | | | Cases | Control | Total | Case-Mean | Control-Mean | | | | | | | | |
| 1 | Schwieler et al. 2015 | Sweden | PS | HC | 23 | 37 | 60 | 36.08 | 23.53 | KA, KYN | CSF | LCMS | Multiple | On Drug | 3 | 10 | (KYN)Ptn > HC***<br>(KA)Ptn > HC** |
| 2 | Huang et al. 2021 | China | SCZ | HC | 195 | 70 | 265 | 35.6 | 39.74 | KA, QA | Serum | LCMS | NR | Mix | 6 | 7.5 | (KA)Ptn < HC***<br>(QA) Ptn < HC**; |
| 3 | Holtze et al. 2012 | Sweden | SCZ | HC | 17 | 33 | 50 | 33.2 | 27.9 | KA | CSF | HPLC | NR | Mix | 3 | 20.5 | (KA)Ptn > HC* |
| 4 | Szymona et al. 2017 | Poland | SCZ | HC | 51 | 45 | 96 | 26.9 | 24.2 | 3HK, KA | Serum | HPLC | NR | Mix | 4 | 11 | (KA)Ptn < HC*<br>(3HK)Ptn > HC |
| 5 | Myint et al. 2011 | South Korea | SCZ | HC | 53 | 48 | 101 | 33.3 | 32.56 | KA, 3HK | Plasma | HPLC | NR | Mix | 5 | 10.5 | (KA)Ptn < HC ***<br>(3HK)Ptn > HC*** |
| 6 | Nilsson et al. 2005 | Sweden | SCZ | HC | 90 | 43 | 133 | 29.9 | 27 | KA | CSF | HPLC | Mix | Mix | 4 | 12 | (KA)Ptn > HC* |
| 7 | Linderholm et al. 2010 | Sweden | SCZ | HC | 16 | 29 | 45 | 36.8 | 25.4 | KYN, KA, TRP | CSF | HPLC | Mix | Mix | 4 | 10.5 | (KYN)Ptn > HC***<br>(KA)Ptn > HC**<br>(TRP) Ptn <HC |
| 8 | Erhardt et al. 2001 | Sweden | SCZ | HC | 28 | 17 | 45 | 27.4 | 27.3 | KA | CSF | HPLC | Mix | Mix | 2 | 21.5 | (KA)Ptn > HC* |
| 9 | Wurfel et al. 2017 | USA | SCZ | HC | 61 | 92 | 153 | 39 | 32.3 | QA, 3HK, KYN, KA, TRP | Serum | LCMS | NR | On Drug | 5 | 20 | (QA)Ptn > HC<br>(3HK)Ptn < HC**<br>(KYN)Ptn =HC<br>(KA)Ptn<HC***<br>(TRP)Ptn < HC |
| 10 | Huang et al. 2020 | China | SCZ | HC | 30 | 34 | 64 | 27.63 | 29.59 | KYN, KA, TRP | Plasma | LCMS | NR | Mix | 5 | 10.5 | (KYN)Ptn > HC<br>(KA)Ptn > HC**<br>(TRP)Ptn < HC |
| 11 | Barry et al. 2009 | Ireland | PS | HC | 34 | 36 | 70 | 37.3 | 33.7 | KYN, KA, TRP | Plasma | HPLC | Multiple | Mix | 4 | 11 | (KYN)Ptn>HC<br>(KA)Ptn<HC<br>(TRP)Ptn < HC |
| 12 | De Picker et al. 2019 | Belgium | PS | HC | 37 | 52 | 89 | 32.4 | 28.5 | 3HK, KYN, KA, TRP, QA | Plasma | LCMS | Mix | Mix | 6 | 9.5 | (3HK)Ptn< HC<br>(KYN)Ptn <HC<br>(KA)Ptn< HC<br>(TRP)Ptn< HC<br>(QA)Ptn< HC |
| 13 | Chiappelli et al. 2018 | USA | PS | HC | 106 | 104 | 210 | 36.3 | 35.4 | KYN, KA | Plasma | HPLC | NR | Mix | 4 | 10 | (KYN)Ptn> HC<br>(KA)Ptn> HC* |

| | | | | | | | | | | | | | | | | | |
|---|---|---|---|---|---|---|---|---|---|---|---|---|---|---|---|---|---|
| 14 | Cathomas et al. 2021 | Switzerland | SCZ | HC | 45 | 19 | 64 | 34 | 32.53 | KA, QA, TRP, 3HK, KYN | Plasma | HPLC | Multiple | Mix | 4 | 8.5 | (KA)Ptn <HC<br>(QA)Ptn<HC<br>(TRP)Ptn HC<br>(3HK)Ptn <HC<br>(KYN)Ptn <HC |
| 15 | Chiappelli et al. 2016 | USA | SCZ | HC | 37 | 38 | 75 | 39.2 | 39.7 | KYN, TRP | Plasma | HPLC | NR | Mix | 4 | 8 | (KYN)Ptn <HC<br>(TRP)Ptn< HC*** |
| 16 | Fazio et al. 2015 | Italy | SCZ | HC | 40 | 84 | 124 | 36.7, 26.9 | 32.8 | 3HA, 3HK, AA, KYN, KA, QA, TRP, XA | Serum | LCMS | Mix | Mix | 4 | 12 | (3HA)Ptn <HC*<br>(3HK)Ptn< HC*<br>(AA)Ptn >HC*<br>(KYN)Ptn > HC<br>(KA)Ptn> HC*<br>(QA)Ptn <HC*<br>(TRP)Ptn> HC<br>(XA)Ptn< HC |
| 17 | Fukushima et al. 2014 | Japan | SCZ | HC | 25 | 27 | 52 | 28.2 | 26.5 | KYN, KA, TRP | Serum | LCMS | Multiple | On Drug | 3 | 17.5 | (KYN)Ptn> HC**<br>(KA)Ptn <HC<br>(TRP)Ptn< HC*** |
| 18 | Kegel et al. 2014 | Sweden | PS | HC | 22 | 26 | 48 | 37.1 | 24.9 | KYN, KA, QA, TRP | CSF | LCMS | First | On Drug | 2 | 11.5 | (KYN)Ptn > HC***<br>(KA)Ptn >HC**<br>(QA)Ptn> HC<br>(TRP)Ptn< HC |
| 19 | Ravikumar et al. 2000 | India | SCZ | HC | 15 | 15 | 30 | NR | NR | KA, QA, TRP | Plasma | HPLC | NR | NR | 3 | 18 | (KA)Ptn> HC**<br>(QA)Ptn> HC**<br>(TRP)Ptn >HC** |
| 20 | Carl et al. 1992 | USA | SCZ | HC | 13 | 13 | 26 | 38.6 | 37.7 | TRP | Plasma | HPLC | Multiple | Mix | 5 | 19.5 | (TRP)Ptn> HC |
| 21 | Cao et al. 2018 | China | SCZ | HC | 208 | 175 | 383 | 37.77 | 39.44 | KYN, TRP | Plasma | LCMS | Mix | Mix | 5 | 10.5 | (KYN)Ptn <HC***<br>(TRP)Ptn <HC*** |
| 22 | Condray et al. 2011 | USA | PS | HC | 25 | 30 | 55 | 22.6 | NR | 3HK, KYN, TRP | Plasma | LCECA | First | Drug Free | 4 | 20.5 | (3HK)Ptn <HC<br>(KYN)Ptn <HC**<br>(TRP)Ptn< HC |
| 23 | Joaquim et al. 2018 | Brazil | SCZ | HC | 28 | 30 | 58 | 26 | 26.2 | KYN, TRP | Plasma | LCMS | First | Drug Free | 3.5 | 16 | (KYN)Ptn <HC**<br>(TRP)Ptn <HC** |
| 24 | Kim et al. 2009 | South Korea | SCZ | HC | 71 | 174 | 245 | 33.9 | 32.49 | KYN , TRP | Plasma | HPLC | Mix | Mix | 5 | 8.5 | (KYN)Ptn< HC<br>(TRP)Ptn< HC** |
| 25 | Lee et al. 2011 | USA | PS | HC | 159 | 55 | 214 | 35.5 | 29 | TRP | Plasma | HPLC | NR | Drug Free | 6 | 9.5 | (TRP)Ptn< HC** |
| 26 | Oxenkrug et al. 2017 | USA | SCZ | HC | 7 | 12 | 19 | NR | NR | 3HK, AA, KYN, KA, TRP | Serum | LCMS | NR | On Drug | 3 | 23 | (3HK)Ptn <HC<br>(AA)Ptn>HC<br>(KYN)Ptn >HC*<br>(KA)Ptn>HC*<br>(TRP)Ptn> HC |

| | | | | | | | | | | | | | | | | | |
|---|---|---|---|---|---|---|---|---|---|---|---|---|---|---|---|---|---|
| 27 | Paredes et al. 2014 | USA | SCZ | HC | 80 | 20 | 100 | 42.5 | 41.1 | KYN | Plasma | LCMS | NR | On Drug | 2 | 17.5 | (KYN)Ptn <HC* |
| 28 | Rao et al. 1990 | Germany | SCZ | HC | 23 | 90 | 113 | 33 | 25 | TRP | Serum | HPLC | NR | Mix | 2 | 21.5 | (TRP)Ptn <HC* |
| 29 | Tortorella et al. 2001 | Italy | SCZ | HC | 11 | 11 | 22 | 24.5 | 29.6 | TRP | Serum | HPLC | NR | On Drug | 4 | 11.5 | (TRP)Ptn <HC*** |
| 30 | van der Heijden et al. 2005 | Netherlands | SCZ | HC | 66 | 73 | 139 | 32.5 | 36.7 | TRP | Plasma | HPLC | NR | Drug Free | 6 | 10.5 | (TRP)Ptn <HC |
| 31 | Leppik et al. 2018 | Estonia | SCZ | HC | 38 | 37 | 75 | 25.4 | 24.8 | KYN, TRP | Serum | LCMS | First | Drug Free | 5.5 | 7 | (KYN)Ptn<HC**<br>(TRP)Ptn <HC** |
| 32 | Potkin et al. 1983 | USA | SCZ | HC | 22 | 18 | 40 | 29.5 | 32.1 | TRP | Plasma | HPLC | Multiple | On Drug | 3.5 | 16 | (TRP)Ptn <HC |
| 33 | Shovestul et al. 2017 | USA | PS | HC | 10 | 10 | 20 | 40.9 | 35.4 | KA, KYN | Serum | HPLC | Multiple | On Drug | 2 | 21.5 | (KA)Ptn>HC<br>(KYN)Ptn< HC |
| 34 | Sathyasaikumar et al. 2011 | USA | SCZ | HC | 15 | 15 | 30 | 50 | 46.7 | KA | Brain tissue | HPLC | NR | On Drug | 2 | 21.5 | (KA)Ptn> HC* |
| 35 | Miller et al. 2006 | USA | SCZ | HC | 12 | 14 | 26 | 43.8 | 48.6 | KA | Brain tissue | HPLC | NR | On Drug | 1 | 25 | (KA)Ptn >HC |
| 36 | Schwarcz et al. 2001 | USA | SCZ | HC | 30 | 31 | 61 | 49.8 | 49.8 | 3HK, KYN, KA | Brain tissue | HPLC | NR | Mix | 3 | 21.5 | (3HK)Ptn> HC<br>(KYN)Ptn> HC<br>(KA)Ptn >HC |
| 37 | Curto et al. 2019 | Italy | PS | HC | 10 | 25 | 35 | 23.3 | 21.6 | 3HK,AA, KYN, KA, QA, TRP, XA | Serum | LCMS | NR | Drug Free | 5 | 20.5 | (3HK)Ptn >HC<br>(AA)Ptn> HC<br>(KYN)Ptn> HC<br>(KA)Ptn >HC<br>(QA)Ptn>HC;<br>(TRP)Ptn>HC*<br>(XA)Ptn <HC* |
| 38 | Oxenkrug et al. 2019 | USA | SCZ | HC | 52 | 52 | 104 | NR | NR | 3HK, KA, XA | Plasma | LCMS | Mix | Mix | 2.5 | 20 | (3HK)Ptn<HC<br>(KA)Ptn<HC*<br>(XA)Ptn <HC* |
| 39 | Wei 1995 | UK | SCZ | HC | 23 | 28 | 51 | 36.5 | 39.9 | TRP | Serum | HPLC | First | Drug Free | 5 | 9 | (TRP)Ptn <HC |
| 40 | Nilsson-Todd et al. 2007 | Sweden | SCZ | HC | 53 | 43 | 96 | 31.1 | NR | KA | CSF | HPLC | First | Drug Free | 5 | 11.5 | (KA)Ptn >HC* |

| | | | | | | | | | | | | | | | | | |
|---|---|---|---|---|---|---|---|---|---|---|---|---|---|---|---|---|---|
| 41 | Zhang et al. 2020 | China | SCZ | HC | 80 | 79 | 159 | 37.5 | 28.85 | KYN, KA, TRP | Serum | LCMS | Mix | Mix | 6 | 14 | (KYN)Ptn< HC**<br>(KA)Ptn <HC**<br>(TRP)Ptn< HC** |
| 42 | Oxenkrug et al. 2016 | USA | SCZ | HC | 6 | 12 | 18 | NR | NR | 3HK, AA ,KYN, KA, TRP | Serum | LCMS | Mix | Mix | 3 | 22 | (3HK)Ptn<HC*<br>(AA)Ptn> HC***<br>(KYN)Ptn>HC<br>(KA)Ptn> HC<br>(TRP)Ptn> HC |
| 43 | Noyan et al. 2021 | Turkey | SCZ | HC | 148 | 210 | 358 | 31.64 | 36.72 | KYN, KA, TRP | Serum | HPLC | NR | Mix | 6 | 12.5 | (KYN)Ptn< C<br>(KA)Ptn> HC<br>(TRP)Ptn< HC** |
| 44 | Yao et al. 2010 | USA | PS | HC | 25 | 30 | 55 | 22.57 | 22.78 | TRP, KYN, 3HK, 3HA,AA | Plasma | LCECA | First | Drug Free | 6 | 9.5 | (TRP)Ptn <HC<br>(KYN)Ptn <HC<br>(3HK)Ptn <HC<br>(3HA)Ptn >HC<br>(AA)Ptn> HC |
| 45 | Okamoto et al. 2021 | Japan | SCZ | HC | 30 | 10 | 40 | 48 | 48 | 3HK, KYN, QA, TRP | Serum | TOFMS | Multiple | On Drug | 8 | 11 | (3HK)Ptn <HC*<br>(KYN)Ptn <HC<br>(TRP)Ptn< C<br>(QA)Ptn <HC |
| 46 | Brundin et al. 2016 | Sweden | PS | HC | 3 | 29 | 32 | 37 | 40 | QA, PA | Plasma | GC-Mass | NR | Mix | 5 | 10.5 | (QA)Ptn < HC<br>(PA)Ptn <HC |
| 47 | Domino 1974 | USA | SCZ | HC | 27 | 36 | 63 | 24 - 46 | 19 - 53 | TRP | Plasma | SPC | Multiple | Drug free | 6 | 12.5 | (TRP)Ptn <HC |
| 48 | Fekkes et al. 1996 | Netherlands | SCZ | HC | 13 | 17 | 30 | 34 | 32 | TRP | Plasma | radioimm unoassay | Mix | Drug free | 6 | 9.5 | (TRP)Ptn <HC* |
| 49 | Rao et al. 1994 | Germany | SCZ | HC | 113 | 34 | 147 | 35, 34 | 24.4 | TRP | Serum | IEC | Multiple | Mix | 5 | 14 | (TRP)Ptn <HC* |
| 50 | Sperner-Unterweger et al. 2002 | Austria | SCZ | HC | 10 | 10 | 20 | 32.3 | NR | TRP, KYN | Serum | HPLC | Multiple | Mix | 5 | 10.5 | (TRP)Ptn < HC<br>(KYN)Ptn > HC |
| 51 | van de Kerkhof et al. 2016 | Netherlands | SCZ | HC | 33 | 75 | 108 | 32.6 | 37.9 | TRP | Plasma | HPLC | Multiple | Mix | 5 | 11.5 | (TRP)Ptn > HC |
| 52 | Kindler et al. 2020 | Australia | SCZ | HC | 96 | 81 | 177 | 35.7 | 31.7 | TRP, KYN, 3HK, QA, KA | Plasma | uHPLC/ GCSM | NR | NR | 6 | 10.5 | (TRP)Ptn > HC**<br>(KYN)Ptn > HC*<br>(3HK)Ptn < HC<br>(QA)Ptn > HC<br>(KA)Ptn > HC** |
| 53 | Koike et al. 2014 | Japan | SCZ | HC | 30 | 38 | 68 | 23.8 | 20.4 | TRP, KYN | Plasma | CE-TOFMS | 0 | Mix | 6 | 10 | (TRP)Ptn > HC<br>(KYN)Ptn > HC |
| 54 | Glassman et al. 2018 | USA | PS | HC | 10 | 10 | 20 | 40.9 | 35.4 | KYN, KA | Serum | HPLC | NR | NR | 5 | 14 | (KYN)Ptn < HC<br>(KA)Ptn > HC |

| | | | | | | | | | | | | | | | | | |
|---|---|---|---|---|---|---|---|---|---|---|---|---|---|---|---|---|---|
| 55 | Steen et al. 2020 | Norway | SCZ | HC | 139 | 68 | 207 | 28.0 | 31.0 | TRP, KYN, 3HK, XA | Plasma | LCECA | Mix | Mix | 5 | 14.5 | (TRP)Ptn < HC<br>(KYN)Ptn < HC<br>(3HK)Ptn> HC<br>(XA)Ptn< HC* |
| 56 | Wu et al. 2019 | China | SCZ | HC | 39 | 60 | 99 | 28 | 31.6 | KYN | Serum | LCMS | NR | NR | | | (KYN)Ptn > HC** |
| 57 | Tang et al. 2009 | China | SCZ | HC | 80 | 80 | 160 | 24.8 | 29.5 | KYN, KA | Serum | HPLC | NR | 0 | | | (KYN)Ptn > HC<br>(KA)Ptn < HC* |
| 58 | Pi et al. 2007 | China | SCZ | HC | 120 | 108 | 228 | 27.4 | 32.2 | TRP, KA | Serum | HPLC | NR | Mix | | | (TRP)Ptn > HC<br>(KA)Ptn < HC |
| 59 | Yang et al. 2016 | China | SCZ | HC | 55 | 33 | 88 | 26.12 | 24.34 | KYN | Serum | ELISA | NR | NR | | | (KYN)Ptn > HC** |
| 60 | Manowitz et al. 1973 | USA | SCZ | HC | 53 | 15 | 68 | NR | NR | TRP | Plasma | HPLC | NR | NR | | | (TRP)Ptn < HC*** |
| 61 | Domingues et al. 2015 | Brazil | SCZ | HC | 35 | 38 | 73 | NR | NR | TRP | Plasma | LCMS | NR | On Drug | 4 | 18.5 | (TRP)Ptn < HC |

* p < 0.05, **p < 0.01, ***p < 0.001; Total quality scores is 10 points; total red point scores are 26 points.

<u>Abbreviations:</u>
TRP: Tryptophan
KYN: Kynurenine
KA: Kynurenic acid
3HK: 3-Hydroxykynurenine
AA: Anthranilic acid
3HA: 3-Hydroxyanthranilic acid
QA: Quinolinic acid
PA: Picolinic acid
XA: Xanthurenic acid
SCZ: Schizophrenia

TRYCATs: Tryptophan catabolites
CSF: Cerebrospinal fluid
SMD: Standardized mean difference
CI: Confidence Interval
O&NS: Oxidative and nitrosative stress
IDO: Indoleamine 2, 3-dioxygenase
CIRS: Compensatory immune-regulatory system
IRS: Immune response system
FEP: First episode psychosis

FES: First episode schizophrenia
MES: Multiple episode schizophrenia
ROS: Reactive oxygen species
5-HT: %-Hydroxy tryptamine
KMO: Kynurenine monooxygenase
BBB: Blood brain Barrier
LAT-1: Large neutral amino acid transporter-1
Phe: Phenylalanine
Tyr: Tyrosine
Leu: Leucine
Ile: Isoleucine
Val: Valine
KNYU: Kynureninase
KATI: Kynurenine aminotransferase I
SSD: Schizophrenia spectrum disorder
HC: Healthy control
DSM: Diagnostic and statistical manual -5
ICD: International Classification of Diseases
SD: Standard deviation
SEM: Standard error
ICS: Immune cofounder scale
HPLC: High performance liquid chromatography
LCMS: Liquid chromatography–mass spectrometry
EDTA: Ethylene diamine tetra acetic acid
HIV: Human immuno-deficiency virus
Ach: Acetylcholine
NE: Norepinephrine
7nAChr: 7 nicotinic acetylcholine receptors
GABA: γ-aminobutyric acid
NMDA: N-Methyl-D-aspartate receptor
LPS: Lipopolysaccharide

**ESF, Table 6.** Results on publication bias.

| Outcome feature sets | Fail safe n | Z Kendall's τ | p | Egger's t test (df) | p | Missing studies (side) |
|---|---|---|---|---|---|---|
| KYN/TRP (Overall) | 4.767 | 0.536 | 0.295 | 1.04 (50) | 0.151 | 0 |
| KYN/TRP (CNS) | 5.638 | 1.959 | 0.025 | 2.23 (3) | 0.051 | 0 |
| KYN/TRP (Plasma) | 0.852 | 0.198 | 0.421 | 0.005 (24) | 0.497 | 0 |
| KYN/TRP (Serum) | 3.802 | 0.301 | 0.381 | 0.42 (19) | 0.338 | 0 |
| (KYN+KA)/TRP (Overall) | 4.444 | 2.103 | 0.017 | 2.44 (59) | 0.008 | 2 (Left) |
| (KYN+KA)/TRP (CNS) | 7.537 | 2.593 | 0.004 | 2.51 (8) | 0.018 | 0 |
| (KYN+KA)/TRP (Plasma) | -0.522 | 0.375 | 0.353 | 0.802 (27) | 0.214 | 3 (Right) |
| (KYN+KA)/TRP (Serum) | 2.919 | 0.817 | 0.206 | 0.879 (20) | 0.194 | 0 |
| (KA+KAT)/(KYN+TRP) (Overall) | 4.622 | 0.698 | 0.242 | 0.164 (60) | 0.434 | 1 (Right) |
| (KA+KAT)/KYN (Overall) | 1.965 | 1.327 | 0.092 | 0.311 (47) | 0.378 | 2 (Right) |
| (3HK+KMO)/KYN (Overall) | -1.747 | 2.426 | 0.007 | 3.50 (39) | 0.0005 | 1 (Left) |
| (3HK+KMO)/KYN (CNS) | -8.118 | 1.315 | 0.094 | 2.00 (40) | 0.057 | 1 (Left) |
| (3HK+KMO)/KYN (Plasma) | 4.077 | -0.157 | 0.172 | 1.51 (17) | 0.073 | 6 (Right) |

| | | | | | | |
|---|---|---|---|---|---|---|
| (3HK+KMO)/KYN (Serum) | -2.269 | 0.360 | 0.359 | 1.47 (14) | 0.081 | 1 (Right) |
| KA (Overall) | 1.906 | 3.187 | 0.0007 | 1.50 (32) | 0.071 | 1 (Left) |
| KA (CNS) | 7.238 | 2.236 | 0.012 | 2.040 (8) | 0.037 | 0 |
| KA (Plasma) | -4.120 | 0.466 | 0.030 | 2.661 (8) | 0.014 | 2 (Right) |
| KA (Serum) | 0.336 | 1.478 | 0.069 | 0.236 (12) | 0.408 | 0 |
| TRP (Overall) | -6.475 | 1.092 | 0.137 | 1.193 (42) | 0.119 | 6 (Right) |
| KYN (Overall) | 0.726 | 2.223 | 0.013 | 3.33 (35) | 0.001 | 0 |
| KYN (CNS) | 8.134 | 0.979 | 0.163 | 1.663 (3) | 0.097 | 1 (Right) |
| KYN (Plasma) | -5.222 | 0.270 | 0.393 | 1.298 (14) | 0.107 | 0 |
| KYN (Serum) | 1.780 | 0.900 | 0.183 | 1.473 (14) | 0.081 | 3 (Left) |

KYN: Kynurenine, TRP: Tryptophan, KA: Kynurenic acid, 3HK: 3-Hydroxykynurenine, KMO: Kynurenine 3-monooxygenase, KAT: Kynurenine aminotransferase.

**ESF, Table 7**: Results of meta-regression

| Variables | No. of Studies | Covariates | 1-sided p-value | Z-Value |
|---|---|---|---|---|
| KYN/TRP | 52 | CNS-Periphery | < 0.001 | -3.22 |
| | | Latitude | 0.032 | 1.85 |
| | 39 | Age | 0.048 | -1.66 |
| KYN+KA/TRP | 61 | CNS-Periphery | < 0.001 | -4.20 |
| | 51 | Female-cases | 0.080 | -1.40 |
| KYN | 37 | CNS-Periphery | < 0.001 | -5.27 |
| | | Latitude | 0.050 | 1.64 |
| | | Studies from Cao | 0.009 | 2.34 |
| | 31 | Age | 0.003 | -2.71 |
| | 33 | Male-cases | 0.019 | -2.07 |
| | | Female-cases | 0.044 | -1.70 |
| | 34 | N-Cases | 0.022 | -2.01 |
| | | Total participant | 0.030 | -1.88 |
| KA | 34 | CNS-Periphery | < 0.001 | -3.11 |
| | 31 | Female-cases | 0.032 | -1.85 |

### (KYN+KA)/TRP

| Study name | Year | Subgroup within study | Std diff in means | Sample size: Patients | Sample size: Control |
|---|---|---|---|---|---|
| Erhardt | 2001 | CNS | 0.611 | 28 | 17 |
| Holtze | 2012 | CNS | 0.157 | 17 | 33 |
| Kegel | 2014 | CNS | 0.704 | 21 | 26 |
| Linderholm | 2012 | CNS | 1.314 | 16 | 29 |
| Miller | 2006 | CNS | 0.679 | 12 | 14 |
| Nilsson-a | 2005 | CNS | 0.485 | 90 | 49 |
| Nilsson-b | 2007 | CNS | 1.235 | 29 | 43 |
| Sathy | 2011 | CNS | 1.699 | 15 | 15 |
| Schwarcz | 2001 | CNS | 0.460 | 30 | 31 |
| Schwieler | 2015 | CNS | 0.663 | 23 | 37 |
| | | CNS | 0.714 | 281 | 294 |
| Barry | 2009 | Plasma | 0.093 | 34 | 36 |
| Cao | 2018 | Plasma | -0.099 | 208 | 175 |
| Carl | 1992 | Plasma | -0.709 | 13 | 13 |
| Cathomas | 2021 | Plasma | -0.148 | 45 | 19 |
| Chiapp-b | 2016 | Plasma | 0.357 | 37 | 38 |
| Chiapp-c | 2018 | Plasma | -0.316 | 106 | 104 |
| Condray | 2011 | Plasma | -0.032 | 25 | 30 |
| De Picker | 2020 | Plasma | -0.314 | 49 | 52 |
| Domingues | 2014 | Plasma | 0.241 | 18 | 38 |
| Domino | 1974 | Plasma | -0.623 | 27 | 36 |
| Fekkes | 1996 | Plasma | 1.250 | 13 | 17 |
| Huang (a) | 2020 | Plasma | 0.477 | 30 | 34 |
| Joaquim | 2018 | Plasma | -0.046 | 28 | 30 |
| Kim | 2009 | Plasma | 0.097 | 71 | 174 |
| Kindler | 2019 | Plasma | -0.081 | 96 | 81 |
| Koike-a | 2014 | Plasma | -0.160 | 18 | 14 |
| Koike-b | 2014 | Plasma | 0.227 | 12 | 24 |
| Lee | 2011 | Plasma | 0.523 | 80 | 55 |
| Manowitz | 1973 | Plasma | 1.597 | 53 | 15 |
| Myint | 2011 | Plasma | -0.668 | 53 | 48 |
| Oxenkrug-b | 2019 | Plasma | -0.328 | 52 | 52 |
| Paredes | 2014 | Plasma | -0.269 | 20 | 20 |
| Potkin | 1983 | Plasma | 0.472 | 22 | 18 |
| Ravikumar | 2000 | Plasma | -0.854 | 15 | 15 |
| Steen | 2019 | Plasma | 0.049 | 139 | 68 |
| Szymona | 2017 | Plasma | -0.573 | 51 | 45 |
| van der | 2005 | Plasma | 0.069 | 66 | 73 |
| van der ke | 2016 | Plasma | -0.089 | 33 | 75 |
| Yao | 2010 | Plasma | -0.103 | 25 | 30 |
| | | Plasma | -0.013 | 1438 | 1429 |
| Curto | 2019 | Serum | -0.070 | 10 | 25 |
| Fazio | 2015 | Serum | 0.138 | 45 | 84 |
| Fukushima | 2014 | Serum | -0.110 | 25 | 27 |
| Glassman | 2018 | Serum | -0.531 | 10 | 10 |
| Huang (b) | 2021 | Serum | -0.659 | 195 | 70 |
| Leppik | 2018 | Serum | 0.141 | 38 | 37 |
| Noyan | 2021 | Serum | 0.490 | 148 | 210 |
| Okamoto | 2021 | Serum | -0.005 | 30 | 10 |
| Oxenkrug-a | 2017 | Serum | 0.810 | 7 | 12 |
| Oxenkrug-c | 2016 | Serum | 0.002 | 6 | 12 |
| Pi | 2007 | Serum | 0.085 | 120 | 108 |
| Rao (a) | 1990 | Serum | 0.726 | 36 | 90 |
| Roa (b) | 1994 | Serum | 0.512 | 57 | 34 |
| Shovestul | 2017 | Serum | -0.050 | 10 | 10 |
| Sperner | 2002 | Serum | 0.713 | 10 | 10 |
| Tang | 2009 | Serum | -0.317 | 80 | 80 |
| Torforella | 2001 | Serum | 1.841 | 11 | 11 |
| Wei | 1995 | Serum | 0.355 | 12 | 28 |
| Wu | 2019 | Serum | 0.226 | 39 | 60 |
| Wurfel | 2017 | Serum | -0.111 | 31 | 92 |
| Yang | 2016 | Serum | 0.607 | 55 | 33 |
| Zhang | 2020 | Serum | -0.102 | 80 | 79 |
| | | Serum | 0.167 | 1054 | 1132 |
| | | Overall | 0.177 | 2772 | 2855 |

Std diff in means and 95% CI

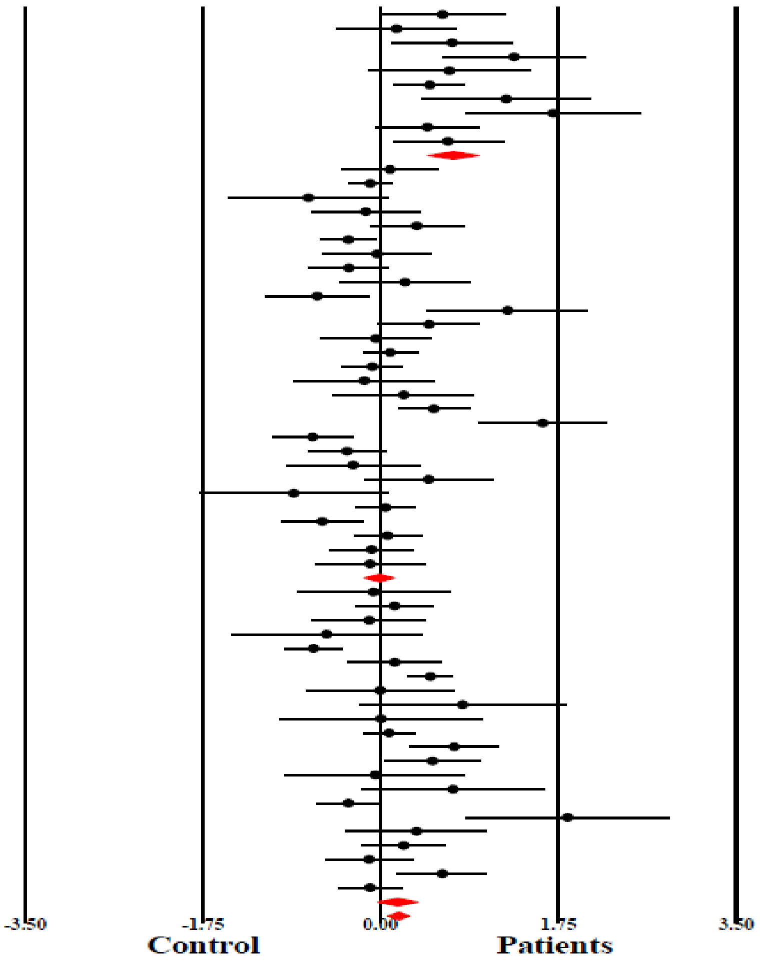

**Almulla et al, 2021**

ESF, Figure 1. Forest plot with results of meta-analysis performed on 61 studies reporting on KA+KYN/TRP ratio, reflecting IDO activity, in schizophrenia.

### (KA+KAT)/(KYN+TRP)

| Study name | Year | Subgroup within study | Std diff in means | Sample size: Patients | Sample size: Control |
|---|---|---|---|---|---|
| Erhardt | 2001 | CNS | 0.611 | 28 | 17 |
| Holtze | 2012 | CNS | 0.157 | 17 | 33 |
| Kegel | 2014 | CNS | 0.028 | 21 | 26 |
| Kindler | 2019 | CNS | 0.642 | 34 | 35 |
| Linderholm | 2012 | CNS | -0.459 | 16 | 29 |
| Miller | 2006 | CNS | -0.263 | 12 | 14 |
| Nilsson-a | 2005 | CNS | 0.485 | 90 | 49 |
| Nilsson-b | 2007 | CNS | 1.235 | 29 | 43 |
| Sathy | 2011 | CNS | 1.699 | 15 | 15 |
| Schwarcz | 2001 | CNS | -0.014 | 30 | 31 |
| Schwieler | 2015 | CNS | -0.198 | 23 | 37 |
| | | CNS | 0.325 | 315 | 329 |
| Barry | 2009 | Plasma | 0.044 | 34 | 36 |
| Cao | 2018 | Plasma | 0.481 | 208 | 175 |
| Carl | 1992 | Plasma | -0.709 | 13 | 13 |
| Cathomas | 2021 | Plasma | 0.058 | 45 | 19 |
| Chiapp-b | 2016 | Plasma | 0.537 | 37 | 38 |
| Chiapp-c | 2018 | Plasma | -0.094 | 106 | 104 |
| Condray | 2011 | Plasma | 0.402 | 25 | 30 |
| De Picker | 2020 | Plasma | -0.111 | 49 | 52 |
| Domingues | 2014 | Plasma | 0.241 | 18 | 38 |
| Domino | 1974 | Plasma | -0.623 | 27 | 36 |
| Fekkes | 1996 | Plasma | 1.250 | 13 | 17 |
| Huang (a) | 2020 | Plasma | 0.147 | 30 | 34 |
| Joaquim | 2018 | Plasma | 0.844 | 28 | 30 |
| Kim | 2009 | Plasma | 0.334 | 71 | 174 |
| Kindler | 2019 | Plasma | 0.218 | 96 | 81 |
| Koike-a | 2014 | Plasma | -0.030 | 18 | 14 |
| Koike-b | 2014 | Plasma | -0.102 | 12 | 24 |
| Lee | 2011 | Plasma | 0.523 | 80 | 55 |
| Manowitz | 1973 | Plasma | 1.597 | 53 | 15 |
| Myint | 2011 | Plasma | -0.668 | 53 | 48 |
| Oxenkrug-b | 2019 | Plasma | -0.328 | 52 | 52 |
| Paredes | 2014 | Plasma | 0.269 | 20 | 20 |
| Potkin | 1983 | Plasma | 0.472 | 22 | 18 |
| Ravikumar | 2000 | Plasma | -0.854 | 15 | 15 |
| Steen | 2019 | Plasma | 0.146 | 139 | 68 |
| Szymona | 2017 | Plasma | -0.573 | 51 | 45 |
| van der | 2005 | Plasma | 0.069 | 66 | 73 |
| van der ke | 2016 | Plasma | -0.089 | 33 | 75 |
| Yao | 2010 | Plasma | 0.523 | 25 | 30 |
| | | Plasma | 0.136 | 1438 | 1429 |
| Curto | 2019 | Serum | -0.480 | 10 | 25 |
| Fazio | 2015 | Serum | -0.038 | 45 | 84 |
| Fukushima | 2014 | Serum | -0.674 | 25 | 27 |
| Glassman | 2018 | Serum | 0.875 | 10 | 10 |
| Huang (b) | 2021 | Serum | -0.659 | 195 | 70 |
| Leppik | 2018 | Serum | 0.437 | 38 | 37 |
| Noyan | 2021 | Serum | 0.689 | 148 | 210 |
| Okamoto | 2021 | Serum | 0.480 | 30 | 10 |
| Oxenkrug-a | 2017 | Serum | -0.380 | 7 | 12 |
| Oxenkrug-c | 2016 | Serum | -0.442 | 6 | 12 |
| Pi | 2007 | Serum | 0.085 | 120 | 108 |
| Rao (a) | 1990 | Serum | 0.726 | 36 | 90 |
| Roa (b) | 1994 | Serum | 0.512 | 57 | 34 |
| Shovestul | 2017 | Serum | 0.328 | 10 | 10 |
| Sperner | 2002 | Serum | 0.039 | 10 | 10 |
| Tang | 2009 | Serum | -0.387 | 80 | 80 |
| Tortorella | 2001 | Serum | 1.841 | 11 | 11 |
| Wei | 1995 | Serum | 0.355 | 12 | 28 |
| Wu | 2019 | Serum | -0.273 | 39 | 60 |
| Wurfel | 2017 | Serum | -0.008 | 31 | 92 |
| Yang | 2016 | Serum | -0.607 | 55 | 33 |
| Zhang | 2020 | Serum | 0.080 | 80 | 79 |
| | | Serum | 0.082 | 1054 | 1132 |
| | | Overall | 0.150 | 2806 | 2890 |

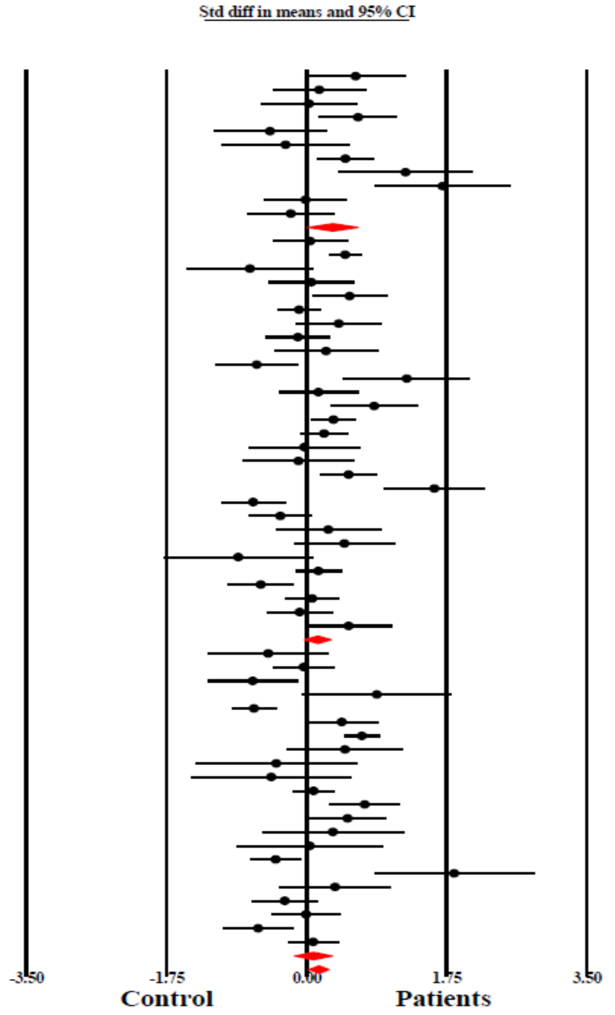

**Almulla et al, 2021**

ESF, Figure 2. Forest plot with results of the meta-analysis performed on 61 studies reporting on (KA+KAT)/(KYN+TRP) ratio, which reflects KAT activity.

### TRP

| Study name | Year | Subgroup within study | Std diff in means | Sample size: Patients | Sample size: Control |
|---|---|---|---|---|---|
| Kegel | 2014 | CNS | -0.368 | 22 | 26 |
| Linderholm | 2012 | CNS | -0.245 | 16 | 29 |
| Schwieler | 2015 | CNS | 0.025 | 23 | 37 |
| | | CNS | -0.179 | 61 | 92 |
| Barry | 2009 | Plasma | -0.328 | 34 | 36 |
| Cao | 2018 | Plasma | -0.382 | 208 | 175 |
| Carl | 1992 | Plasma | 0.709 | 13 | 13 |
| Cathomas | 2021 | Plasma | -0.089 | 45 | 19 |
| Chiapp-b | 2016 | Plasma | -0.895 | 37 | 38 |
| Condray | 2011 | Plasma | -0.369 | 25 | 30 |
| De Picker | 2020 | Plasma | -0.089 | 49 | 52 |
| Domingues | 2014 | Plasma | -0.241 | 18 | 38 |
| Domino | 1974 | Plasma | 0.623 | 27 | 36 |
| Fekkes | 1996 | Plasma | -1.250 | 13 | 17 |
| Huang (a) | 2020 | Plasma | -0.137 | 30 | 34 |
| Joaquim | 2018 | Plasma | -0.798 | 28 | 30 |
| Kim | 2009 | Plasma | -0.431 | 71 | 174 |
| Kindler | 2019 | Plasma | -0.799 | 96 | 81 |
| Koike-a | 2014 | Plasma | 0.190 | 18 | 14 |
| Koike-b | 2014 | Plasma | -0.125 | 12 | 24 |
| Lee | 2011 | Plasma | -0.523 | 80 | 55 |
| Manowitz | 1973 | Plasma | -1.597 | 53 | 15 |
| Potkin | 1983 | Plasma | -0.472 | 22 | 18 |
| Ravikumar | 2000 | Plasma | 3.004 | 15 | 15 |
| Steen | 2019 | Plasma | -0.196 | 139 | 68 |
| van der | 2005 | Plasma | -0.069 | 66 | 73 |
| van der ke | 2016 | Plasma | 0.089 | 33 | 75 |
| Yao | 2010 | Plasma | -0.420 | 25 | 30 |
| | | Plasma | -0.256 | 1156 | 1160 |
| Curto | 2019 | Serum | 1.102 | 10 | 25 |
| Fazio | 2015 | Serum | 0.141 | 45 | 84 |
| Fukushima | 2014 | Serum | 0.986 | 25 | 27 |
| Leppik | 2018 | Serum | -0.578 | 38 | 37 |
| Noyan | 2021 | Serum | -0.433 | 148 | 210 |
| Okamoto | 2021 | Serum | -0.475 | 30 | 10 |
| Oxenkrug-a | 2017 | Serum | 0.492 | 7 | 12 |
| Oxenkrug-c | 2016 | Serum | 0.716 | 6 | 12 |
| Pi | 2007 | Serum | 0.211 | 120 | 108 |
| Rao (a) | 1990 | Serum | -0.726 | 36 | 90 |
| Roa (b) | 1994 | Serum | -0.512 | 57 | 34 |
| Sperner | 2002 | Serum | -0.752 | 10 | 10 |
| Tortorella | 2001 | Serum | -1.841 | 11 | 11 |
| Wei | 1995 | Serum | -0.355 | 12 | 28 |
| Wu | 2019 | Serum | 0.071 | 39 | 60 |
| Wurfel | 2017 | Serum | -0.227 | 31 | 92 |
| Zhang | 2020 | Serum | -0.536 | 80 | 79 |
| | | Serum | -0.164 | 704 | 929 |
| | | Overall | -0.213 | 1921 | 2181 |

Std diff in means and 95% CI

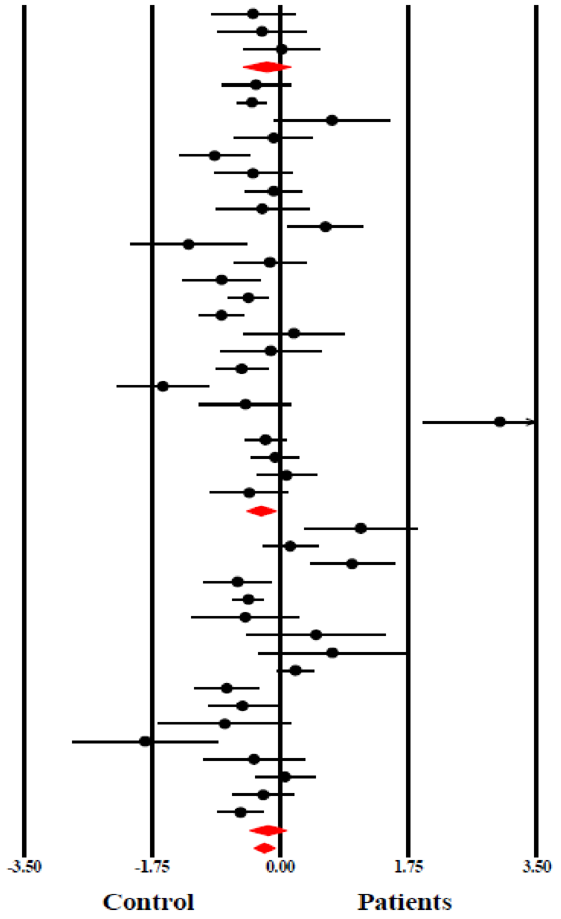

**Almulla et al, 2021**

ESF, Figure 3. Forest plot with results of meta-analysis performed on 44 studies reporting tryptophan levels in schizophrenia

## KA

| Study name | Year | Subgroup within study | Std diff in means | Sample size: Patients | Sample size: Control |
|---|---|---|---|---|---|
| Erhardt | 2001 | CNS | 0.611 | 28 | 17 |
| Holtze | 2012 | CNS | 0.157 | 17 | 33 |
| Kegel | 2014 | CNS | 0.730 | 19 | 26 |
| Linderholm | 2012 | CNS | 1.039 | 16 | 29 |
| Miller | 2006 | CNS | 0.416 | 12 | 14 |
| Nilsson-a | 2005 | CNS | 0.485 | 90 | 49 |
| Nilsson-b | 2007 | CNS | 1.235 | 29 | 43 |
| Sathy | 2011 | CNS | 1.699 | 15 | 15 |
| Schwarcz | 2001 | CNS | 0.446 | 30 | 31 |
| Schwieler | 2015 | CNS | 0.723 | 23 | 37 |
| | | CNS | 0.676 | 279 | 294 |
| Barry | 2009 | Plasma | -0.123 | 34 | 36 |
| Cathomas | 2021 | Plasma | -0.224 | 45 | 19 |
| Chiapp-c | 2018 | Plasma | -0.410 | 106 | 104 |
| De Picker | 2020 | Plasma | -0.726 | 49 | 52 |
| Huang (a) | 2020 | Plasma | 0.798 | 30 | 34 |
| Kindler | 2019 | Plasma | -0.594 | 96 | 81 |
| Myint | 2011 | Plasma | -0.668 | 53 | 48 |
| Oxenkrug-b | 2019 | Plasma | -0.328 | 52 | 52 |
| Ravikumar | 2000 | Plasma | 1.295 | 15 | 15 |
| Szymona | 2017 | Plasma | -0.573 | 51 | 45 |
| | | Plasma | -0.225 | 531 | 486 |
| Curto | 2019 | Serum | 0.277 | 10 | 25 |
| Fazio | 2015 | Serum | 0.292 | 45 | 84 |
| Fukushima | 2014 | Serum | -0.190 | 25 | 27 |
| Glassman | 2018 | Serum | 0.344 | 10 | 10 |
| Huang (b) | 2021 | Serum | -0.659 | 195 | 70 |
| Noyan | 2021 | Serum | 1.337 | 148 | 210 |
| Oxenkrug-a | 2017 | Serum | 1.137 | 7 | 12 |
| Oxenkrug-c | 2016 | Serum | 0.056 | 6 | 12 |
| Pi | 2007 | Serum | -0.040 | 120 | 108 |
| Shovestul | 2017 | Serum | 0.277 | 10 | 10 |
| Tang | 2009 | Serum | -0.704 | 80 | 80 |
| Wu | 2019 | Serum | -0.141 | 39 | 60 |
| Wurfel | 2017 | Serum | -0.406 | 31 | 92 |
| Zhang | 2020 | Serum | -0.570 | 80 | 79 |
| | | Serum | 0.042 | 806 | 879 |
| | | Overall | 0.288 | 1615 | 1659 |

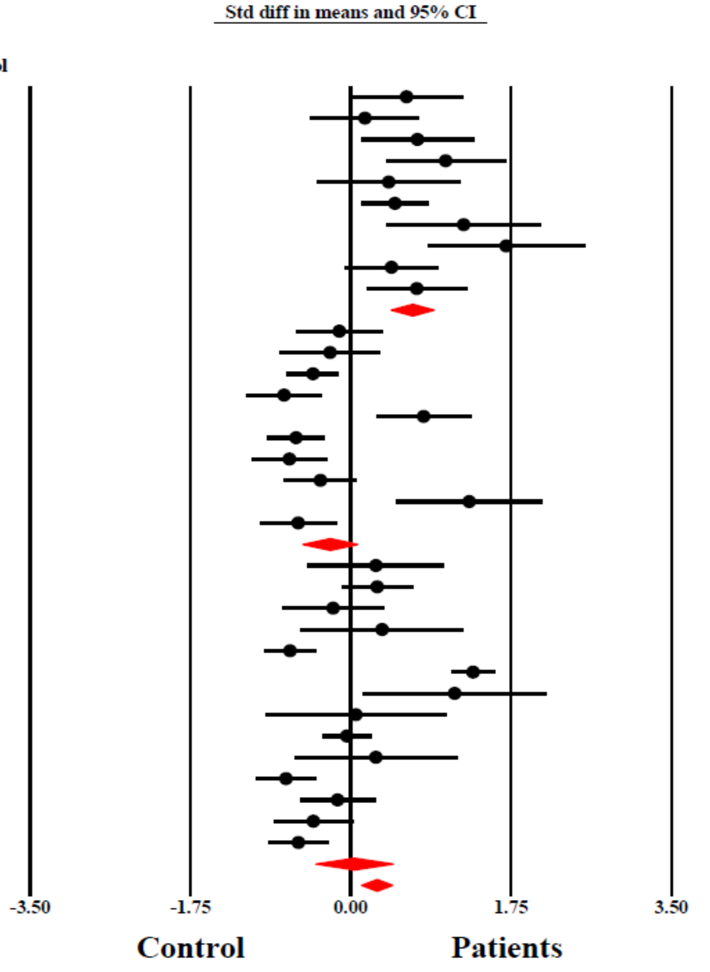

**Almulla et al, 2021**

ESF, Figure 4. Forest plot with results of meta-analysis performed on 34 studies reporting kynurenic acid levels in schizophrenia
